# A Clinical Atlas for Phenome-Wide Association Study of Obstructive Sleep Apnea Across Electronic Health Record Domains

**DOI:** 10.64898/2026.10.20.26366073

**Authors:** Nathanael H. Hwang, Soniya Mishra, M. Brandon Westover, Robert J. Thomas, Umakanth Katwa, Kendra A. Becker, Dennis Hwang, Matthew T. Klimper, Jessica Jara, Damien R. Stevens, Matthew K.P. Gratton, Diego R. Mazzotti, Haoqi Sun

**Affiliations:** Department of Neurology, Beth Israel Deaconess Medical Center, Harvard Medical School, Boston, MA, United States of America; Department of Sleep Medicine, Kaiser Permanente Southern California, Fontana, CA, United States of America; Division of Medical Informatics, Department of Internal Medicine, University of Kansas Medical Center, Kansas City, KS, United States of America; Department of Biostatistics and Data Science, University of Kansas Medical Center, Kansas City, KS, United States of America; Department of Neurology, Stanford University, Stanford, CA, United States of America; Division of Sleep Medicine, Beth Israel Deaconess Medical Center, Harvard Medical School, Boston, MA, United States of America; Boston Children’s Hospital, Harvard Medical School, Boston, MA, United States of America; Division of Pulmonary Critical Care and Sleep Medicine, Department of Internal Medicine, University of Kansas Medical Center, Kansas City, KS, United States of America; Department of Psychiatry and Behavioral Sciences, University of Kansas Medical Center, Kansas City, KS, United States of America

## Abstract

Obstructive sleep apnea (OSA) is a heterogeneous disorder whose associations across the entire phenome remain incompletely characterized by hypothesis-driven outcome-specific analyses. We constructed a multidomain atlas for phenome-wide association study of OSA in 222,109 adults undergoing diagnostic sleep testing in Kaiser Permanente Southern California. Using apnea-hypopnea index, we examined cross-sectional associations across diagnoses, laboratory tests, medications, procedures, healthcare utilization, behaviors, and symptoms; associations with incident diseases; and data-driven cross-domain phenotypes. After full adjustment, 169 prevalent phecodes and 16 incident phecodes remained Bonferroni-significant. Positive associations converged most consistently on cardiometabolic diseases, including hypertension, impaired glucose metabolism, hyperlipidemia, obesity, and heart failure, with concordant laboratory, medication, and cardiac-measurement signals. Multidimensional physiologic, symptom, and comorbidity phenotypes showed heterogeneous disease incidence after adjustment, with patterns not aligned with apnea-hypopnea index severity alone. These findings define the broad clinical landscape of OSA and provide a comprehensive atlas for replication and hypothesis generation.

## INTRODUCTION

Obstructive sleep apnea (OSA) is a prevalent condition affecting nearly one billion adults aged 30 to 69 years worldwide^1^. OSA is a heterogeneous condition, with substantial variation in symptom expression, comorbidity, and clinical consequences^2–5^. Presence of OSA has been associated with a broad range of conditions including obesity, hypertension, atrial fibrillation, stroke, type 2 diabetes, and depression^6–11^. However, these relationships have largely been evaluated in hypothesis-driven disease-specific analyses, with limited systematic comparison across the clinical phenome. Conditions not hypothesized a priori remain largely untested, null associations may be underreported, and differences in study populations, outcome definitions, and analytic approaches complicate comparisons across conditions. The published record is therefore incompletely ascertained by hypothesis rather than by phenome.

The phenome-wide association study (PheWAS) design was developed in the field genetics to address a structurally analogous circumstance^12,13^. As a phenotypic complement to genome-wide association studies, PheWAS reverses the conventional association framework: rather than evaluating many exposures against a single outcome, a single exposure is systematically evaluated against a broad range of phenotypes ascertainable from the electronic health record (EHR), using a standard analytic framework and correction for multiple testing^14^. Anticipated and unanticipated associations are therefore estimated under identical assumptions not driven by prespecified hypotheses, and phenotypes without association are recorded rather than omitted. The PheWAS framework has subsequently been extended to non-genetic clinical exposures, sometimes termed phenome-wide disease association studies (PheDAS), providing an opportunity to systematically characterize the clinical disease landscape associated with a defined exposure^15,16^, and this framework has been previously applied to sleep disorders^17–19^. The same phenome-wide concept can be extended to other EHR domains, including laboratory measurements (LabWAS)^20^ and medications (MedWAS)^21^, and can be further adapted to novel analyses of healthcare utilization (UtilWAS) and health behaviors (BehWAS), each capturing dimensions of health and disease not represented by diagnostic codes alone.

Here, we applied this framework to construct a comprehensive clinical phenome-wide atlas of OSA among adults that underwent diagnostic sleep testing at Kaiser Permanente Southern California, a large integrated healthcare system with near-comprehensive capture of clinical data and stable longitudinal membership. We first evaluated associations between conventional apnea-hypopnea index (AHI)-defined OSA severity and the clinical phenome, including diagnoses and the other domains captured before the index diagnostic sleep study. We then evaluated incident diagnoses following sleep study to characterize prospective disease associations. We also performed age-sex-stratified subset analysis. Finally, recognizing that AHI alone incompletely captures OSA heterogeneity^3,4,22,23^, we derived multidimensional OSA phenotypes additionally incorporating sleep-study physiology and symptom characteristics and evaluated whether these phenotypes identify distinct patterns of subsequent disease incidence beyond conventional AHI-based severity. Complete summary statistics are provided in the *Supplementary Atlas* (Excel workbook) and through an interactive web portal at https://www.osaatlas.com/, creating a resource for hypothesis generation and further investigation of the clinical landscape of OSA.

## METHODS

### Study design and participants

We conducted a retrospective cohort study of adults (≥18 years old) undergoing diagnostic polysomnography or home sleep apnea testing in Kaiser Permanente Southern California during 2016–2025. The diagnostic sleep study date served as the index date. For patients with multiple studies, the earliest eligible study was selected. We excluded patients with missing age, sex, or body mass index (BMI), fewer than two pre-index diagnostic encounters, less than one year of pre-index diagnostic history, or an OSA diagnosis before index, yielding 222,109 participants. The excluded participants were 26.7% (n=80,820) of the original cohort. This study was conducted under the Human Sleep Project protocols, approved by the BIDMC Institutional Review Board (#2024P000804/#2022P000417) with waiver of informed consent for this retrospective study. Full cohort definitions and data-processing methods are provided in the *Supplementary Methods*.

### OSA exposure, covariates, and statistical analysis

OSA severity was defined from the apnea AHI (4% hypopnea criterion) as normal (<5 events/h), mild (5 to <15), moderate (15 to <30), or severe (≥30). The primary contrast was AHI ≥5 versus <5. Model m1 was unadjusted, m2 added age, sex, race/ethnicity, and index year, and m3 additionally included smoking and neighborhood income, with observation-window and encounter-rate terms included as specified for each analysis. The fully adjusted m4 model additionally included BMI. Binary, continuous, and rate outcomes were analyzed using logistic, rank-inverse-normal linear, and offset Poisson models, respectively. Cox proportional hazards models were used for incident outcomes, with participants followed until the outcome occurred, death, disenrollment, or the end of follow-up. Missing smoking status and neighborhood income were retained as explicit unknown/missing categories rather than imputed. Continuous measurements and questionnaire items were analyzed among participants with observed values for each feature; unanswered questionnaire items were not treated as negative responses. Domain-specific target measures, denominators, minimum feature thresholds, and additional missing-data rules are detailed in the *Supplementary Methods*. Overall, missingness was handled using prespecified domain-specific approaches without general imputation. Bonferroni correction at a family-wise α of 0.05 was the primary significance criterion within prespecified analysis families. Benjamini-Hochberg false discovery rate (FDR) results were retained as secondary findings.

### Phase 1: Cross-sectional association atlas (prevalent PheDAS)

Baseline diagnoses were mapped to phecodes^24^ (v1.2) for prevalence PheWAS (also labeled as phenome-wide disease association study or PheDAS). Parallel wide-association studies evaluated laboratory values and ordering (LabWAS), outpatient medication classes (MedWAS), procedure rates (ProcWAS), echocardiographic and spirometric measurements (CRMWAS), healthcare utilization (UtilWAS), health behaviors (BehWAS), and pre-index patient-reported symptoms (QWAS). Continuous outcomes were rank-inverse-normal transformed, rather than log-transformed, before linear regression; coefficients therefore represent differences on an approximately standard-normal scale. Continuous covariates were separately z-standardized before modeling. Most longitudinal EHR domains were summarized over 1- and 5-year pre-index windows. The main figures show the 5-year window, while results for both windows are reported in the *Supplementary Atlas*.

### Phase 2: Incident disease (incidence PheDAS)

Incident phecodes were defined by a second distinct post-index occurrence among patients with no baseline record of that phecode or any related phecode among patients with no baseline record of that phecode or a predefined related phecode. Follow-up was censored at last contact (administrative end or disenrollment) or death. Death was treated as a competing event. We fit cause-specific Cox models for 1,013 outcomes with at least 20 events and estimated crude Aalen-Johansen cumulative incidence.

### Phase 3: Cross-domain phenotype construction and incidence

We fit separate three-class finite-mixture models: a Gaussian latent profile model for 10 physiologic indicators, a mixed continuous/categorical latent class model for 40 symptom indicators, and a Bernoulli latent class model for 172 chronic-disease comorbidity indicators. Class selection considered information criteria, entropy, class size, likelihood replication, nondegeneracy, and split-half stability, as detailed in the Supplementary Methods. Relative entropy was 0.925, 0.792, and 0.917, and split-half adjusted Rand index was 0.966, 0.926, and 0.981 for physiology, symptoms, and comorbidity, respectively. Physiologic and symptom models used full-information maximum likelihood for incomplete indicators under a missing-at-random assumption. The AHI was excluded from model input, but the physiologic model retained two AHI-derived indicators (T90/AHI ratio and the supine-to-nonsupine AHI ratio); AHI was also used to describe the resulting phenotypes. For the 70,880 participants included in the analysis and scored in all three domains, each domain-severity score was calculated as the posterior-probability-weighted average of class ranks (0, 1, and 2), with classes clinically ordered from lower to higher burden within each domain. Splitting each score at its shared-cohort median created eight 2×2×2 cross-domain phenotype strata. These strata are regions of continuous burden gradients and not necessarily discrete biological subpopulations, and the cohort-derived cut points are not clinical thresholds.

Prospective phenotype analyses used a hierarchical design across 368 eligible incident phecodes. A 7-df omnibus Wald test first assessed whether the eight strata differed, and outcomes with omnibus P<0.05 entered eight one-versus-rest Cox contrasts. The adjusted m4 estimates used ridge penalization (0.01, with 0.1 fallback), and multiplicity was controlled within each focal-phenotype family. Selected unadjusted Aalen-Johansen curves show absolute risk with death competing. Adjusted hazard ratios and the complete gated atlas are reported in the *Supplementary Atlas (PheDAS Incidence tab)*.

### OSA treatment and PAP-adherence landmark analysis

Post-index OSA treatment was not included as a covariate in the primary OSA–disease analyses because it may lie on the pathway between OSA and subsequent outcomes. In a separate observational analysis restricted to participants with OSA, PAP adherence was classified at a 180-day post-index landmark using the first 90-day usage summary after device setup, only when that entire window fell between index and landmark. The 90-day usage categories were: ≥4, 2 to <4, and 0 to <2 hours/night, never-started, and unknown. Patients had to be alive, enrolled, and free of the outcome diagnoses at the landmark. Cause-specific Cox models adjusted for baseline covariates and OSA severity, using ≥4 hours/night as the reference, across 40 selected incidence phecodes and 10 negative-control phecodes.

### Data availability

Complete aggregate summary statistics for all WAS domains and Phase 3 phenotype-incidence associations are provided in the *Supplementary Atlas (Excel workbook)* and through https://www.osaatlas.com/. No individual-level data are included.

## RESULTS

### Study cohort

The final analytic cohort *(Table 1)* included 222,109 adults: 161,491 (72.7%) with OSA (AHI ≥5) and 60,618 (27.3%) with No OSA (AHI <5). Mean age was 51.3±15.2 years, mean AHI was 19.9±22.3 events/h, mean BMI was 33.3±7.9 kg/m^2^, and 53.2% were male. The cohort included substantial racial and ethnic diversity with 39.6% Hispanic, 36.8% White, 10.4% Asian, and 8.3% Black participants. Mild, moderate, and severe OSA accounted for 67,040 (30.2%), 44,957 (20.2%), and 49,494 (22.3%) participants, demonstrating broad distribution of OSA severities. Unless otherwise specified, results are from the m4 level of adjustment for AHI ≥5 versus <5, and significance denotes Bonferroni-adjusted P<0.05.

**Table 1.** Baseline characteristics of the analytic cohort, overall and by OSA status.

| Characteristic | Overall | OSA<br>(AHI ≥5) | No OSA<br>(AHI <5) |
| --- | --- | --- | --- |
| Participants, n | 222,109 | 161,491 | 60,618 |
| Age, years, mean (SD) | 51.3 (15.2) | 53.0 (14.6) | 46.7 (15.8) |
| AHI, events/h, mean (SD) | 19.9 (22.3) | 26.6 (22.9) | 2.2 (1.4) |
| BMI, kg/m<sup>2</sup>, mean (SD) | 33.3 (7.9) | 34.4 (7.9) | 30.3 (7.1) |
| Male, n (%) | 118,251 (53.2%) | 93,780 (58.1%) | 24,471 (40.4%) |
| Charlson Comorbidity Index, mean (SD) | 1.48 (2.02) | 1.58 (2.08) | 1.20 (1.83) |
| **Race/ethnicity, n (%)** |  |  |  |
| Hispanic | 87,928 (39.6%) | 64,553 (40.0%) | 23,375 (38.6%) |
| White | 81,732 (36.8%) | 58,926 (36.5%) | 22,806 (37.6%) |
| Asian | 23,109 (10.4%) | 16,748 (10.4%) | 6,361 (10.5%) |
| Black | 18,538 (8.3%) | 13,454 (8.3%) | 5,084 (8.4%) |
| Pacific Islander | 1,726 (0.8%) | 1,384 (0.9%) | 342 (0.6%) |
| American Indian or Alaska Native | 474 (0.2%) | 357 (0.2%) | 117 (0.2%) |
| Multiple | 923 (0.4%) | 620 (0.4%) | 303 (0.5%) |
| Other | 3,205 (1.4%) | 2,227 (1.4%) | 978 (1.6%) |
| Unknown | 4,474 (2.0%) | 3,222 (2.0%) | 1,252 (2.1%) |
| **Smoking status, n (%)** |  |  |  |
| Never | 155,330 (69.9%) | 109,836 (68.0%) | 45,494 (75.1%) |
| Former | 40,840 (18.4%) | 31,857 (19.7%) | 8,983 (14.8%) |
| Current | 13,988 (6.3%) | 10,617 (6.6%) | 3,371 (5.6%) |
| Unknown | 11,951 (5.4%) | 9,181 (5.7%) | 2,770 (4.6%) |
| **Neighborhood income group, n (%)** |  |  |  |
| High (≥\$80,001) | 97,944 (44.1%) | 70,681 (43.8%) | 27,263 (45.0%) |
| Middle (\$45,001–\$80,000) | 58,777 (26.5%) | 42,994 (26.6%) | 15,783 (26.0%) |
| Low (≤\$45,000) | 8,179 (3.7%) | 5,969 (3.7%) | 2,210 (3.6%) |
| Missing | 57,209 (25.8%) | 41,847 (25.9%) | 15,362 (25.3%) |
| **Neighborhood education, n (%)** |  |  |  |
| 0%–50% with ≥12 years of education | 3,491 (1.6%) | 2,497 (1.5%) | 994 (1.6%) |
| 51%–75% with ≥12 years of education | 42,205 (19.0%) | 31,142 (19.3%) | 11,063 (18.3%) |
| 76%–100% with ≥12 years of education | 119,176 (53.7%) | 85,986 (53.2%) | 33,190 (54.8%) |
| Missing neighborhood education | 57,237 (25.8%) | 41,866 (25.9%) | 15,371 (25.4%) |

### Phase 1: Prevalence and multi-domain association studies

#### Disease prevalence landscape (PheDAS)

The fully adjusted prevalence atlas showed a selective positive disease profile within a much larger set of inverse associations (*Figure 1)*. Of 1,780 estimable phecodes, 169 met Bonferroni correction: 29 had ORs above 1 and 140 had ORs below 1. The positive associations centered around cardiometabolic conditions including: essential hypertension (OR 1.32, 95% CI 1.29–1.36), elevated blood pressure without diagnosed hypertension (OR 1.22, 95% CI 1.19–1.25), obesity (OR 1.20, 95% CI 1.17–1.23), and renal dialysis (OR 1.72, 95% CI 1.44–2.05). In contrast, other anemias (OR 0.79, 95% CI 0.77–0.81), constipation (OR 0.78, 95% CI 0.76–0.81), and palpitations (OR 0.82, 95% CI 0.80–0.84) were inversely associated (all P<0.0001). The full disease landscape, complete adjustment tiers, and severity contrasts are provided in the Supplementary Atlas (PheDAS Prevalence tab).

**Figure 1.**
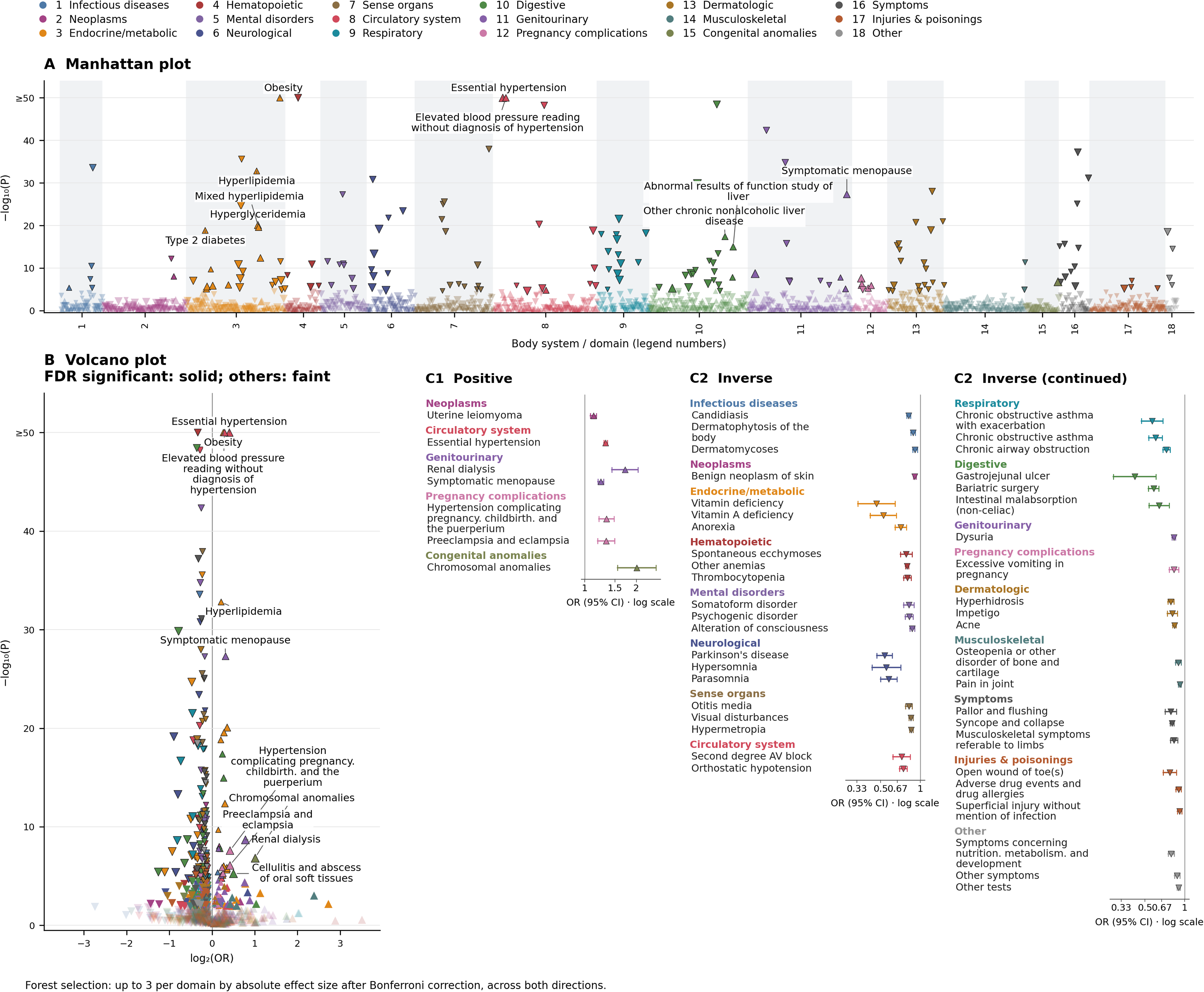
PheDAS Prevalence. A, Manhattan plot; B, volcano plot; C1/C2, positive/inverse forests. Fully adjusted (m4) AHI ≥5 versus <5 associations for 1,779 phecodes, excluding sleep apnea and subcodes. Complete results are provided in the **Atlas, PheDAS Prevalence tab**. Colors denote body systems/domains; triangles indicate effect direction, symbol size reflects effect magnitude, and dark outlines denote Bonferroni significance. Manhattan plots label up to ten positive Bonferroni-significant associations by smallest *P* value; volcano plots label up to five by largest effect size, with solid symbols indicating FDR significance (*q*<0.05). Inverse associations are not labeled. Forest plots show up to three Bonferroni-significant features per body system/domain, selected by absolute effect size; whiskers indicate 95% CIs. Ratio measures use a null of 1 and β measures a null of 0. The −log10(*P*) axis is capped at 50. C1 includes 7 of 29 positive Bonferroni-significant associations; forest plots are selected summaries and are not exhaustive.

#### Other WAS domains: concordant metabolic signals

*Figures 2–6* present Manhattan, volcano, and forest plots for LabWAS, MedWAS, ProcWAS, UtilWAS, and QWAS, respectively; *Figure 7* presents BehWAS forest plots. Most plots display estimates using the 5-year window, if applicable. Both 1- and 5-year estimates are available in the *Supplementary Atlas* (LabWAS, MedWAS, ProcWAS, CRMWAS, UtilWAS, BehWAS, and QWAS tabs).

**Figure 2A.**
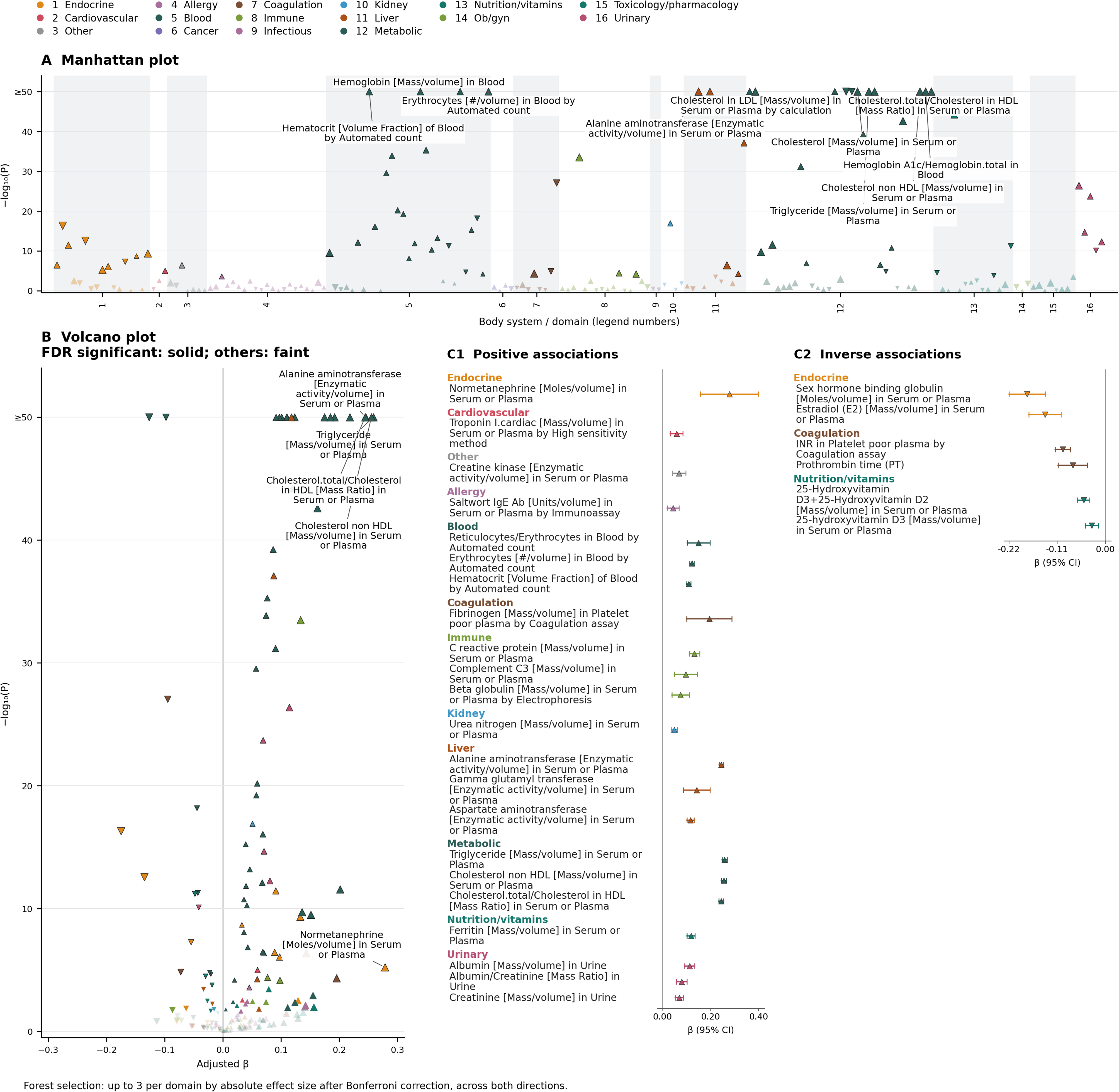
Five-year LabWAS of median laboratory values. A, Manhattan plot; B, volcano plot; C1/C2, positive/inverse forest plots. Analyses include 185 laboratory measures with estimable fully adjusted (m4) rank-inverse-normal β estimates among measured patients, comparing AHI ≥5 versus <5. Complete 1- and 5-year results are provided in the **Atlas, LabWAS tab**. Colors denote laboratory categories; triangles indicate effect direction, symbol size reflects effect magnitude, and dark outlines denote Bonferroni significance. Manhattan plots label up to ten positive Bonferroni-significant associations by smallest *P* value; volcano plots label up to five by largest effect size, with solid symbols indicating FDR significance (*q*<0.05). Inverse associations are not labeled. Forest plots show up to three Bonferroni-significant features per category, selected by absolute effect size; whiskers indicate 95% CIs. The −log10(*P*) axis is capped at 50.

**Figure 2B.**
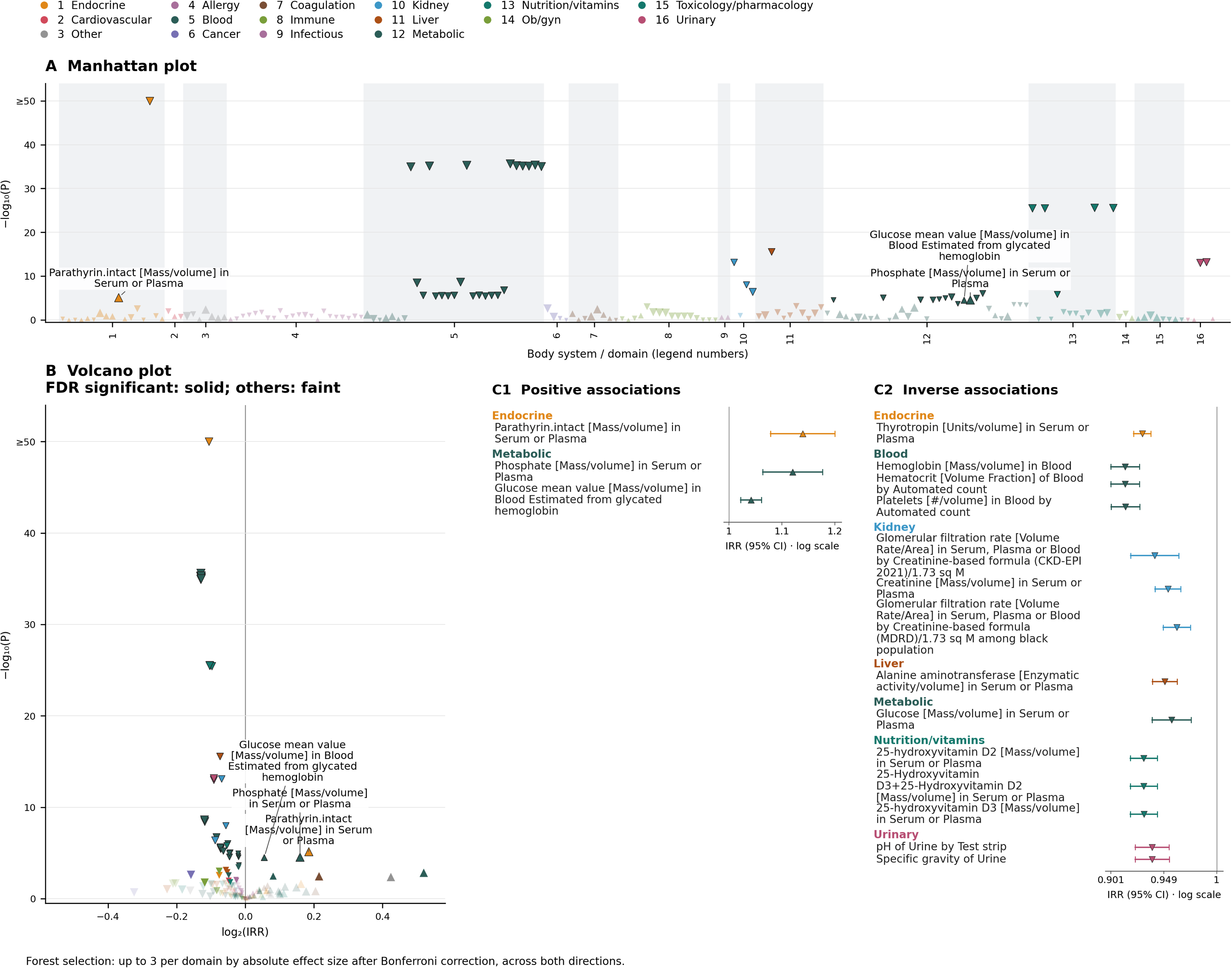
Five-year LabWAS laboratory ordering rates. A, Manhattan plot; B, volcano plot; C1/C2, positive/inverse forests. Five-year LabWAS of laboratory ordering rates. Analyses include 186 laboratory measures with estimable fully adjusted (m4) IRRs among patients with laboratory observations, comparing AHI ≥5 versus <5. Complete 1- and 5-year results are provided in the **Atlas, LabWAS tab**. Colors denote laboratory categories; triangles indicate effect direction, symbol size reflects effect magnitude, and dark outlines denote Bonferroni significance. Manhattan plots label up to ten positive Bonferroni-significant associations by smallest *P* value; volcano plots label up to five by largest effect size, with solid symbols indicating FDR significance (*q*<0.05). Inverse associations are not labeled. Forest plots show up to three Bonferroni-significant features per category, selected by absolute effect size; whiskers indicate 95% CIs. The −log10(*P*) axis is capped at 50.

**Figure 2C.**
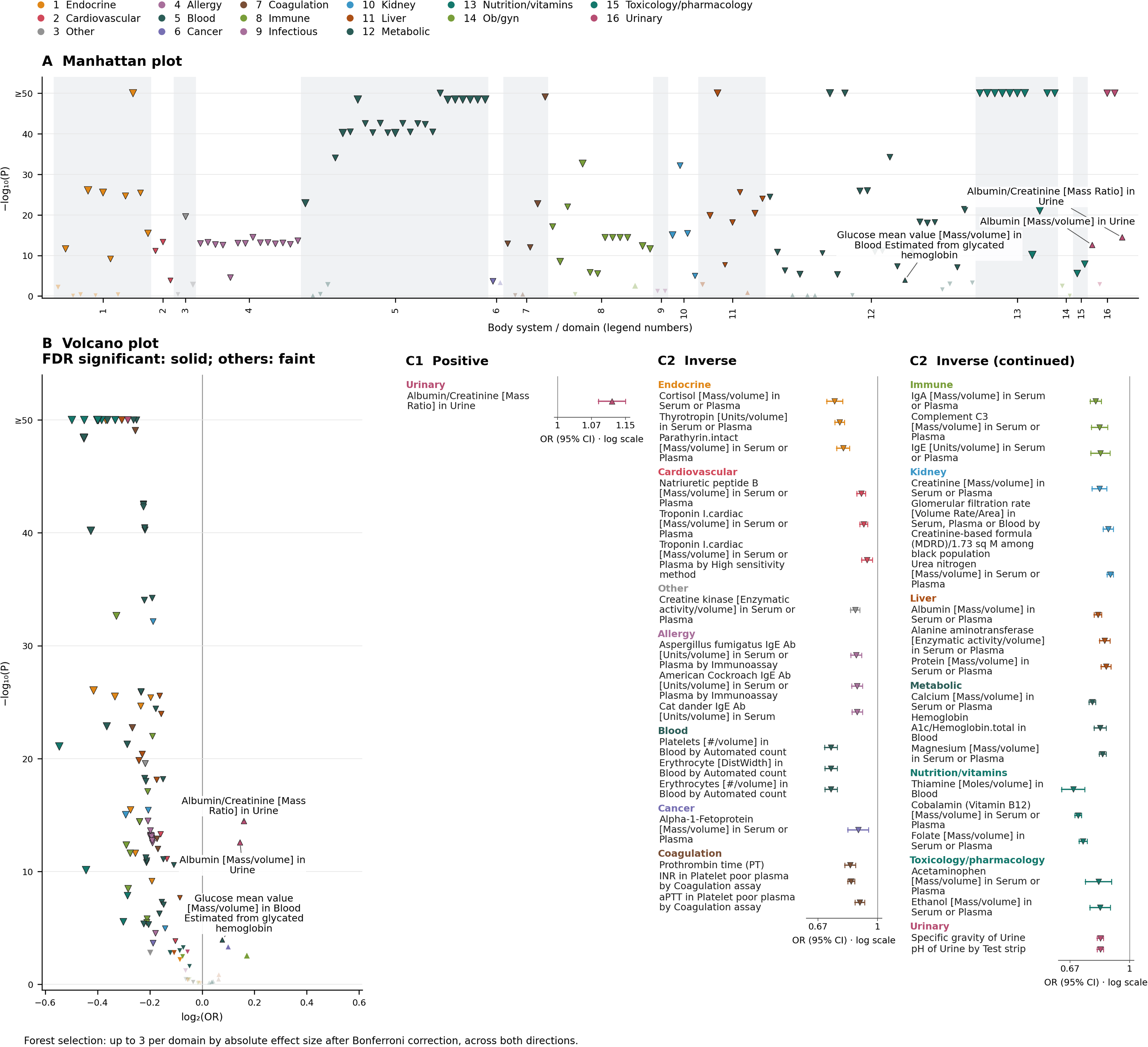
Five-year LabWAS any-order probability. A, Manhattan plot; B, volcano plot; C1/C2, positive/inverse forests. Five-year LabWAS of laboratory ordering propensity. Analyses include 143 laboratory measures with estimable fully adjusted (m4) ORs for any laboratory order; four non-estimable measures were omitted. All contrasts compare AHI ≥5 versus <5. Complete 1- and 5-year results are provided in the **Atlas, LabWAS tab**. Colors denote laboratory categories; triangles indicate effect direction, symbol size reflects effect magnitude, and dark outlines denote Bonferroni significance. Manhattan plots label up to ten positive Bonferroni-significant associations by smallest *P* value; volcano plots label up to five by largest effect size, with solid symbols indicating FDR significance (*q*<0.05). Inverse associations are not labeled. Forest plots show up to three Bonferroni-significant features per category, selected by absolute effect size; whiskers indicate 95% CIs. The −log10(*P*) axis is capped at 50.

For LabWAS, median values were Bonferroni-significant for 64 of 148 measures at 1 year and 74 of 185 at 5 years *(Figure 2).* Generally, laboratory values extended the cardiometabolic pattern. At 5 years, OSA was associated with higher triglycerides (β 0.259, 95% CI 0.249–0.269), alanine aminotransferase (β 0.25, 95% CI 0.24–0.26), non-HDL cholesterol (β 0.26, 95% CI 0.25–0.27), and hemoglobin A1c (β 0.19, 95% CI 0.18–0.19), and lower HDL cholesterol (β −0.10, 95% CI −0.11 to −0.09; all P<0.0001). Laboratory ordering showed a different pattern. Ordering rates were significant for 26 of 149 analytes at 1 year and 47 of 186 at 5 years, while any-order probability was significant for 103 of 122 and 114 of 143 estimable outcomes, respectively. Most significant ordering associations, however, were inverse. Thus, the metabolic differences among measured participants coexisted with lower testing rates for many analytes.

For MedWAS, medication fills provided a corresponding treatment profile. Cardiovascular classes were positively associated with OSA at both 1 and 5 years, including antihypertensive combinations (OR 1.51 and 1.47, respectively), angiotensin II receptor antagonists (1.37 and 1.35), calcium-channel blockers (1.33 and 1.26), thiazide or thiazide-like diuretics (1.39 and 1.32), and HMG-CoA reductase inhibitors (1.21 and 1.20). Despite this concentration of positive associations among cardiovascular therapies, the broader MedWAS landscape was again predominantly inverse: among the 72 of 177 medication classes that were Bonferroni-significant at 1 year and 96 of 218 at 5 years, 50 (69.4%) and 74 (77.1%), respectively, were inversely associated with OSA. *Figure 3* shows the 5-year results.

**Figure 3.**
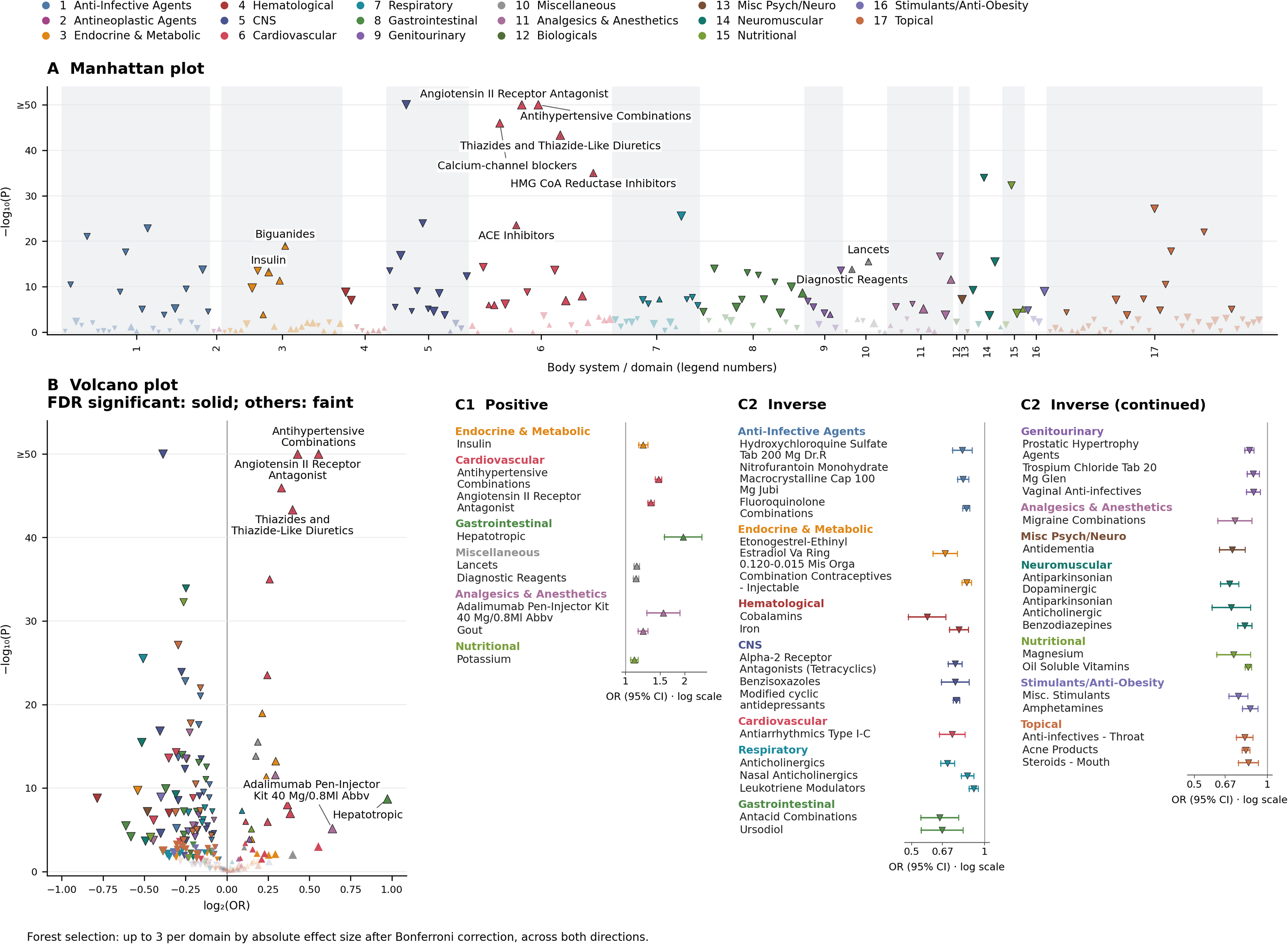
Five-year MedWAS. A, Manhattan plot; B, volcano plot; C1/C2, positive/inverse forests. Five-year MedWAS of 218 medication classes using fully adjusted (m4) AHI ≥5 versus <5 associations. Complete results are provided in the **Atlas, MedWAS tab**. Colors denote body systems/domains; triangles indicate effect direction, symbol size reflects effect magnitude, and dark outlines denote Bonferroni significance. Manhattan plots label up to ten positive Bonferroni-significant associations by smallest *P* value; volcano plots label up to five by largest effect size, with solid symbols indicating FDR significance (*q*<0.05). Inverse associations are not labeled. Forest plots show up to three Bonferroni-significant features per domain, selected by absolute effect size; whiskers indicate 95% CIs. The −log10(*P*) axis is capped at 50.

For ProcWAS, the pattern was overwhelmingly inverse. All 10 Bonferroni-significant procedure-rate associations at 1 year and 16 of 17 at 5 years had IRRs below 1, among 25 and 67 procedures tested, respectively. Examples included extended ambulatory electrocardiographic monitoring over 48 hours to 7 days at 1 year (IRR 0.67, 95% CI 0.60–0.75), and pre/post-bronchodilator spirometry (IRR 0.74, 95% CI 0.70–0.80) and external ECG monitoring up to 48 hours (IRR 0.73, 95% CI 0.68–0.79) at 5 years. Thus, ProcWAS did not identify a coherent positive association between OSA and procedure utilization in any clinical domain, including cardiovascular care. *Figure 4* presents the 5-year results.

**Figure 4.**
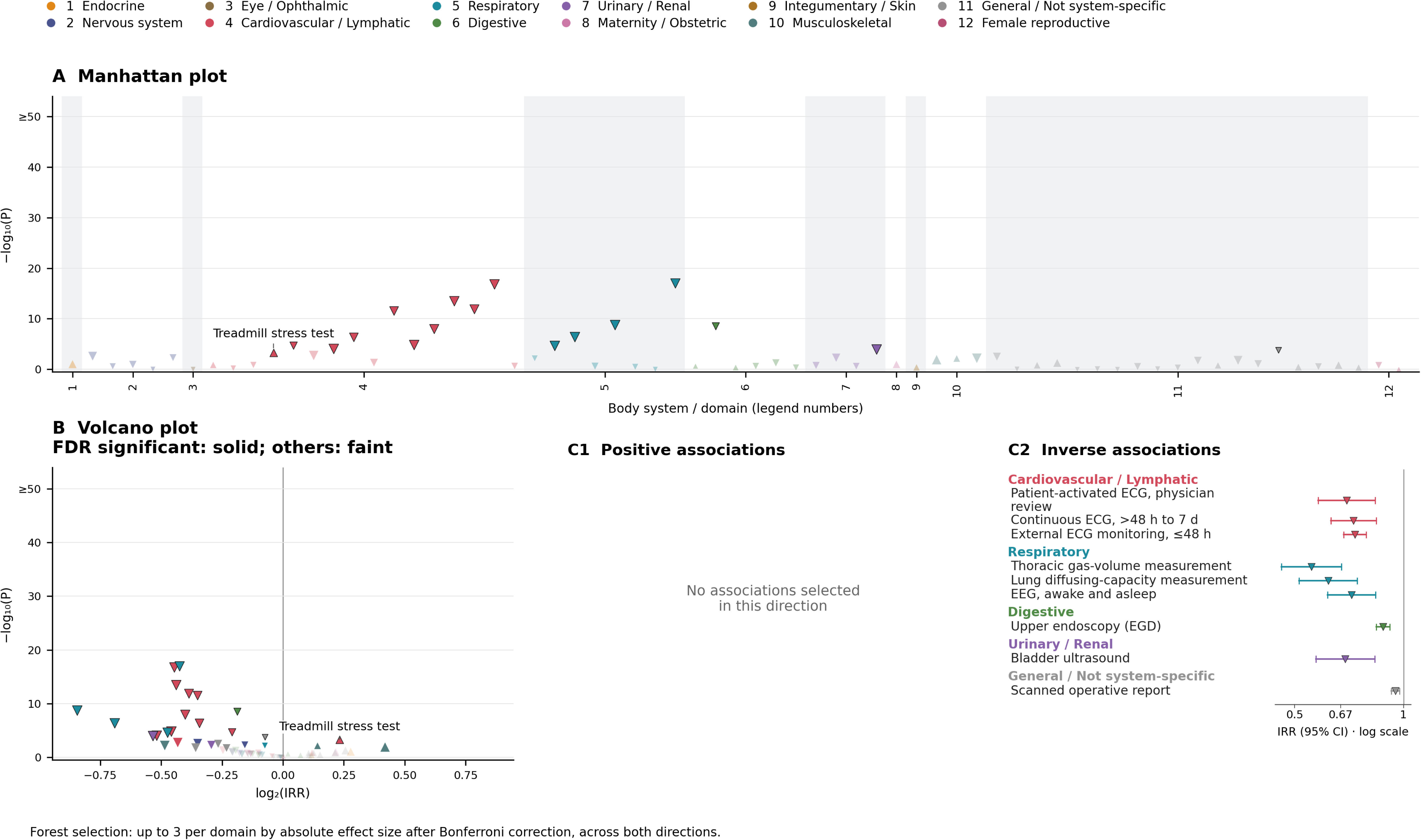
Five-year ProcWAS. A, Manhattan plot; B, volcano plot; C1/C2, positive/inverse forests. Five-year ProcWAS of 67 procedures using fully adjusted (m4) AHI ≥5 versus <5 associations. Complete results are provided in the **Atlas, ProcWAS tab**. Colors denote body systems/domains; triangles indicate effect direction, symbol size reflects effect magnitude, and dark outlines denote Bonferroni significance. Manhattan plots label up to ten positive Bonferroni-significant associations by smallest *P* value; volcano plots label up to five by largest effect size, with solid symbols indicating FDR significance (*q*<0.05). Inverse associations are not labeled. Forest plots show up to three Bonferroni-significant features per domain, selected by absolute effect size; whiskers indicate 95% CIs. The −log10(*P*) axis is capped at 50.

For UtilWAS, *Table 2* shows that healthcare use varied by care setting rather than increasing uniformly with OSA severity. Outpatient encounter rates were counter-intuitively lower in severe OSA than in no OSA: 9.0 ± 9.1 versus 7.2 ± 8.0 encounters/person-year in the 5-year window. Emergency department (ED) and urgent-care rates were also lower. Inpatient encounter rates were higher in severe OSA than in no OSA, increasing from 0.08 to 0.11 encounters/person-year at 1 year and from 0.06 to 0.07 at 5 years, with longer mean hospital length of stay of 4.7 versus 4.0 days/admission at 1 year and 4.5 versus 4.1 days at 5 years. All encounter-rate comparisons across severity groups had unadjusted omnibus Welch ANOVA P<0.0001.

**Table 2.**
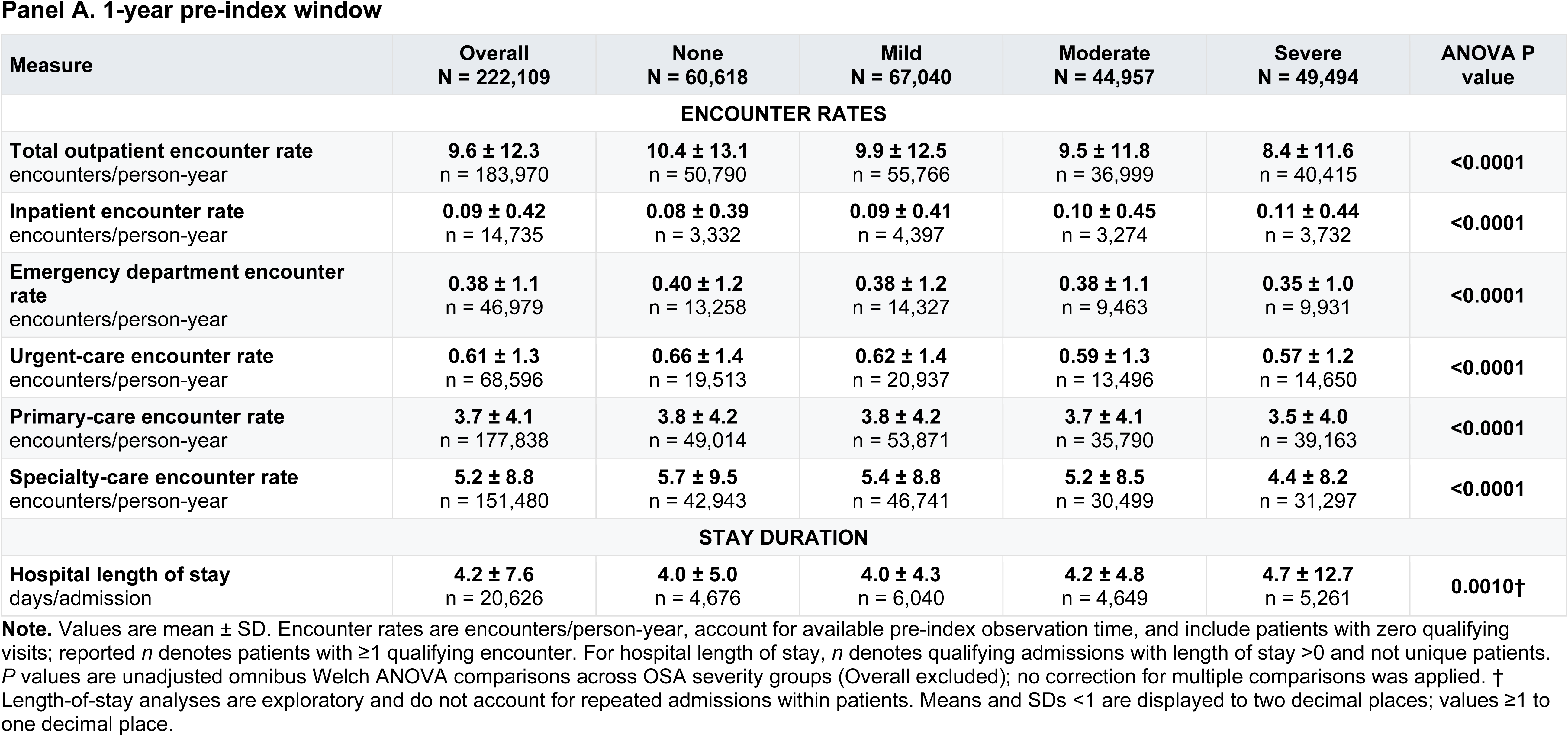
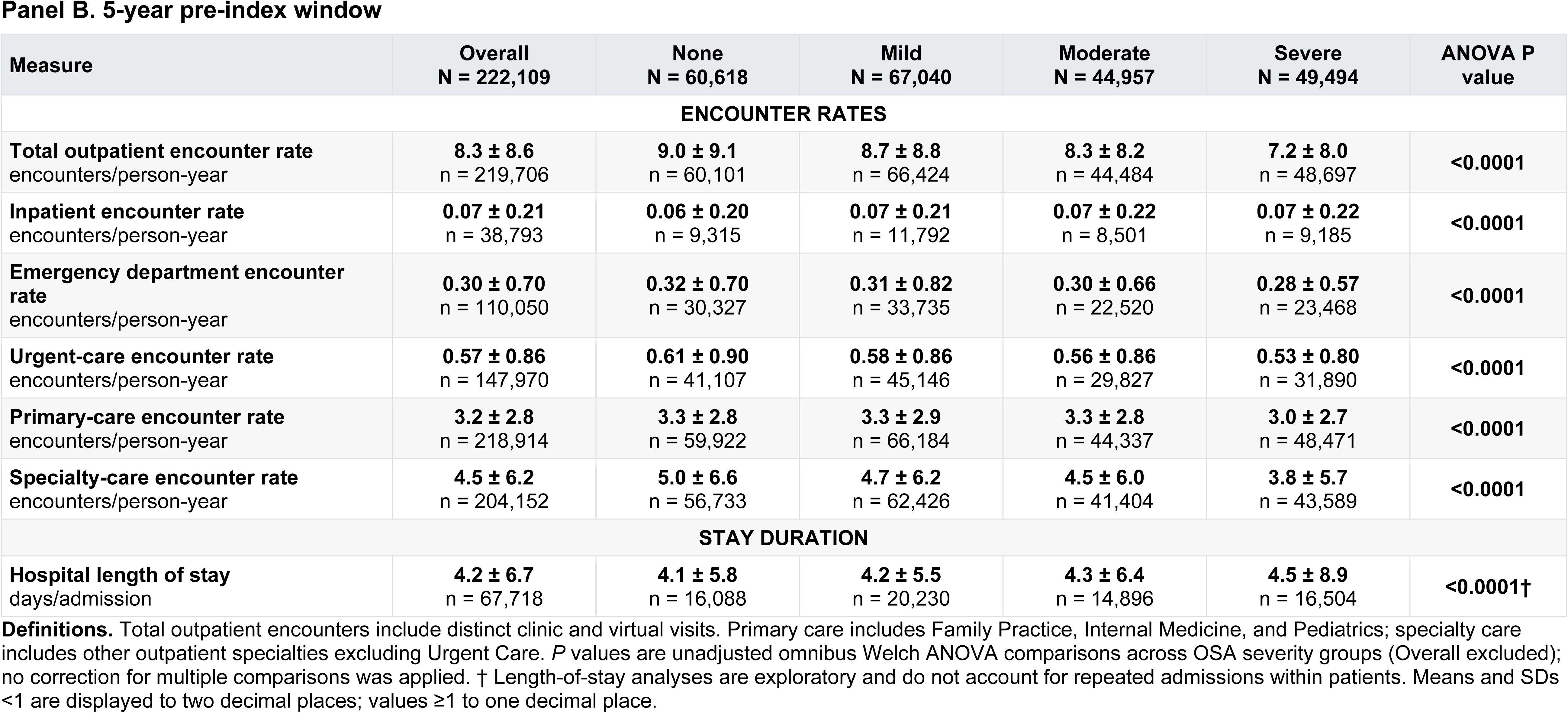
Descriptive healthcare-utilization profile by OSA severity.

ED and outpatient encounter associated diagnoses again revealed a cardiometabolic signature. ED-associated diagnoses are shown in *Figure 5A–B.* At 5 years, 8 of 59 ED-primary and 20 of 145 ED-secondary diagnosis-presence associations were Bonferroni-significant. Essential hypertension was positively associated with OSA as an ED-primary diagnosis (OR 1.21, 95% CI 1.11–1.33) and as an ED-secondary diagnosis (OR 1.12, 95% CI 1.09–1.16). Outpatient-associated diagnoses are shown in *Figure 5C.* At 5 years, 33 of 248 physician-primary diagnosis-presence associations were Bonferroni-significant, including essential hypertension (OR 1.29, 95% CI 1.24–1.33), type 2 diabetes (OR 1.21, 95% CI 1.16–1.27), and heart failure (OR 1.54, 95% CI 1.24–1.91). Across all diagnosis settings, 39 of 205 associations at 1 year and 86 of 685 at 5 years met Bonferroni correction. Specialty encounter-rate associations were Bonferroni-significant for 27 of 80 specialties at 1 year and 36 of 102 at 5 years, mostly inversely associated with *OSA (Figure 5D–E).* Neurology rates were lower at 1 year (IRR 0.67, 95% CI 0.64–0.70) and 5 years (IRR 0.72, 95% CI 0.69–0.75), as were family-medicine (IRR 0.93 in both windows) and pulmonary-clinic encounters (IRR 0.77 and 0.75).

**Figure 5.**
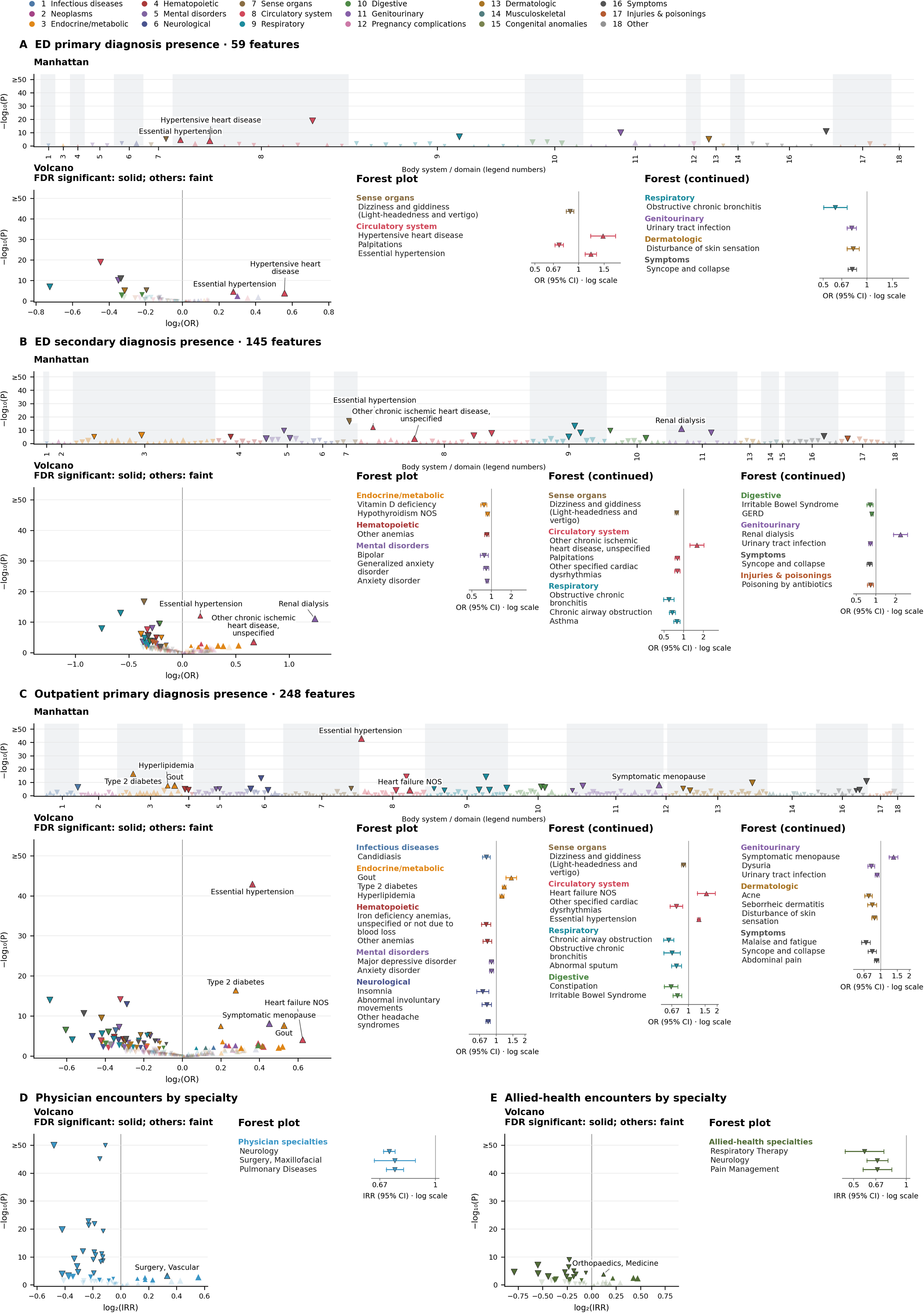
Five-year UtilWAS. A–C combine Manhattan, volcano, and forest plots; D and E show volcano and forest plots only. Five-year UtilWAS using fully adjusted (m4) AHI ≥5 versus <5 associations. **A/B**, ED primary/secondary diagnosis presence; **C**, diagnoses associated with outpatient physician encounters; **D/E**, physician/allied-health encounter rates by specialty. Diagnosis-presence ORs use the full cohort, whereas specialty IRRs reflect encounters per person-time. Colors denote phecode body systems or source specialty groups; triangles indicate effect direction, symbol size reflects effect magnitude, and dark outlines denote Bonferroni significance. Manhattan plots label up to ten positive Bonferroni-significant associations; volcano plots label up to five by largest effect size, with solid symbols indicating FDR significance (*q*<0.05). Forest plots show up to three Bonferroni-significant features per domain; whiskers indicate 95% CIs. Complete results are provided in the **Atlas, UtilWAS tab**.

For QWAS, patient-reported features distinguished the expected breathing-related presentation of OSA from less uniform differences in daytime symptoms. At time of diagnostic evaluation, 33 of 83 binary and 34 of 49 continuous questionnaire features were Bonferroni-significant *(Figure 6).* Snoring on the STOP screen (OR 2.67, 95% CI 2.57–2.77), chief complaint of snoring/apnea (OR 2.45, 95% CI 2.34–2.57), observed apnea (OR 2.17, 95% CI 2.09–2.26), and nocturnal gasping (OR 1.58, 95% CI 1.51–1.66) showed positive associations. OSA was also associated with greater sleepiness (ESS: OR 1.35, 95% CI 1.30–1.41; β 0.154, 95% CI 0.137–0.171), greater insomnia burden (ISI: OR 1.11, 95% CI 1.07–1.16; β 0.078, 95% CI 0.060–0.095), and worse function (FOSQ-10 β −0.042, 95% CI −0.059 to −0.025). PHQ-2 was not significant for the primary binary OSA contrast.

**Figure 6.**
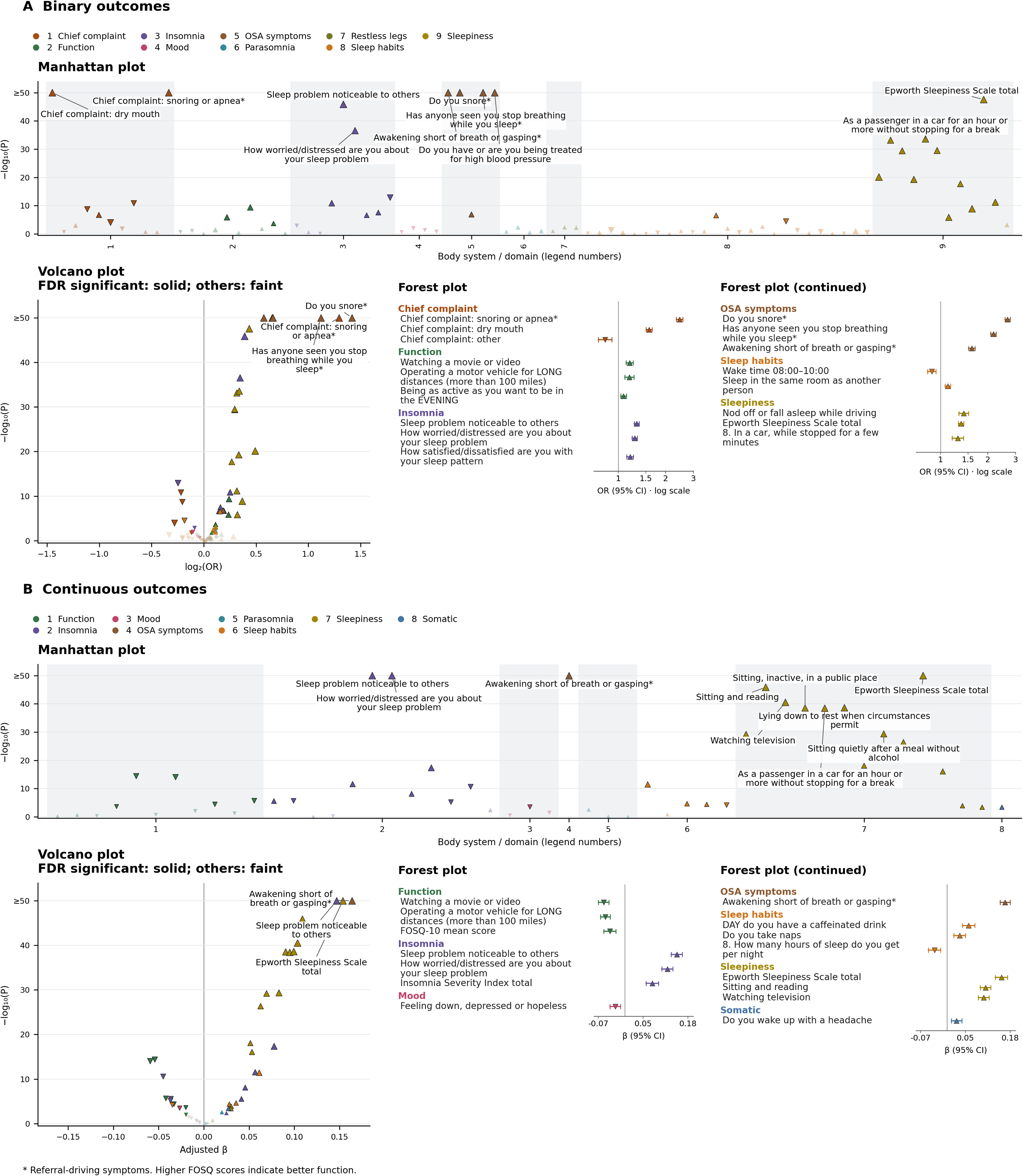
QWAS. A, binary outcomes; B, continuous outcomes. Each section combines Manhattan, volcano, and forest plots. QWAS of 83 binary and 49 continuous questionnaire features using fully adjusted (m4) AHI ≥5 versus <5 associations. Complete results are provided in the **Atlas, QWAS tab**. Colors denote questionnaire domains; triangles indicate effect direction, symbol size reflects effect magnitude, and dark outlines denote Bonferroni significance. Manhattan plots label up to ten positive Bonferroni-significant associations; volcano plots label up to three by largest effect size, with solid symbols indicating FDR significance (*q*<0.05). Inverse associations are not labeled. Forest plots show up to three Bonferroni-significant features per domain, selected by absolute effect size; whiskers indicate 95% CIs. The −log10(*P*) axis is capped at 50.

For BehWAS, five of eight health behavior outcomes were Bonferroni-significant at 1 year and four of eight at 5 years *(Figure 7).* At both windows, OSA was associated with slightly greater medication adherence among users (adherent OR 1.05; PDC β 0.027), but lower influenza vaccination (receipt OR 0.94; count β −0.031 and −0.032 at 1 and 5 years, respectively). Medical-leave filing was not associated with OSA. Among lifetime measures, OSA was associated with lower odds of missed appointments (OR 0.92, 95% CI 0.88–0.95) and higher odds of an off-work order (OR 1.09, 95% CI 1.02–1.16), without an association with total off-work days. *Figure 7* shows 5-year and lifetime results.

**Figure 7.**
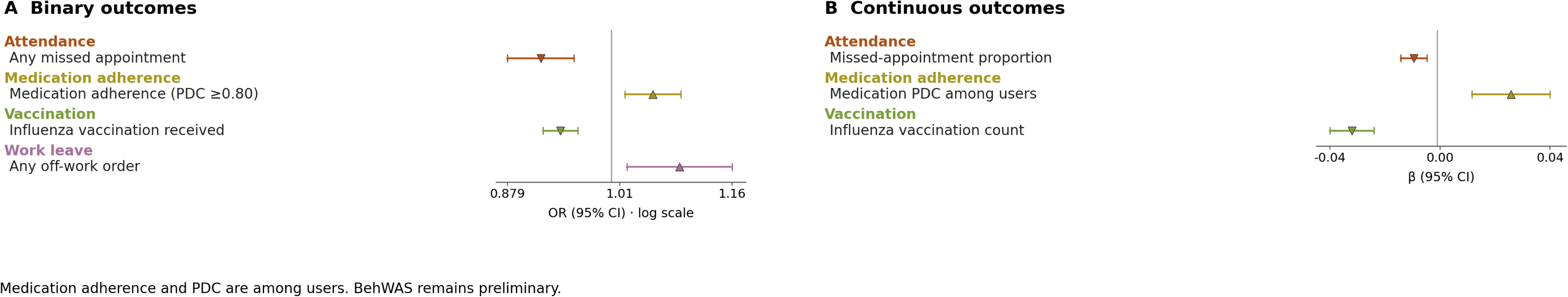
BehWAS. A, binary-outcome forest plot (OR); B, continuous-outcome forest plot (β). BehWAS using fully adjusted (m4) AHI ≥5 versus <5 associations. Five-year estimates are shown for windowed outcomes and lifetime estimates for undated outcomes. Colors denote medication adherence, vaccination, attendance, and work-leave domains. Forest plots show selected Bonferroni-significant associations, with up to three features per domain ranked by absolute effect size; whiskers indicate 95% CIs. Medication adherence and PDC analyses are restricted to hypertension and hypercholesterolemia medication users. Complete results are provided in the **Atlas, BehWAS tab**.

For CRMWAS, cardiac measurements supplied a physiologic counterpart to the disease and medication findings centered around cardiovascular burden. Among participants with echocardiography within the 5-year pre-index window, 12 of 22 measurements were Bonferroni-significant. OSA was associated with greater left-ventricular mass (β 0.111, 95% CI 0.086–0.136), higher E/e′ (β 0.083, 95% CI 0.054–0.113), lower septal e′ velocity (β −0.112, 95% CI −0.140 to −0.083), and lower ejection fraction across its available measures (β −0.082 to −0.115). These differences described structural and both systolic and diastolic functional cardiac changes with OSA.

CRMWAS also evaluated spirometry results which could be suggestive of greater airflow obstruction with OSA but without a suggestion of restrictive ventilatory defect. 10 of 14 comparisons were Bonferroni-significant after full adjustment, with higher pre- and post-bronchodilator FEV1/FVC (β 0.100, 95% CI 0.054–0.145, and β 0.115, 95% CI 0.063–0.167, respectively) and FEV1 (β 0.086, 95% CI 0.049–0.123, and β 0.088, 95% CI 0.045–0.131); the FVC comparison was not significant. FEV1/FVC also increased across OSA severity categories before and after bronchodilator use (ordinal trend, both P<0.0001), with moderate and severe OSA contrasts remaining Bonferroni-significant.

#### Patterns across WAS domains, adjustments, and stratification by OSA severity

Across WAS domains, the positive associations formed a consistent cardiometabolic pattern: hypertension and diabetes diagnoses were accompanied by greater use of cardiovascular medications, higher triglycerides and hemoglobin A1c, lower HDL cholesterol, and echocardiographic differences in ventricular structure and function. This concordance contrasted with predominantly inverse associations across domains including diseases, laboratory ordering, medication classes overall, procedures, and specialty encounters.

The prevalence association landscape changed substantially across adjustment tiers. Bonferroni-significant associations numbered 548 in m1, 303 after demographic and index-year adjustment in m2, 318 after additional adjustment for smoking, income, and observation-window terms in m3, and 169 after BMI was added in m4. For cardiometabolic outcomes, effect estimates generally were attenuated at both the demographic and BMI adjustment steps. Across m1–m4, ORs were 2.31, 1.81, 1.80, and 1.32 for essential hypertension; 2.00, 1.58, 1.57, and 1.15 for type 2 diabetes; and 1.84, 1.34, 1.33, and 1.17 for heart failure NOS. All three remained Bonferroni-significant in m4. Similar reductions after BMI adjustment were evident in other domains. In the 5-year window, the number of Bonferroni-significant median laboratory-value associations declined from 100 in m3 to 74 in m4, medication-fill associations from 118 to 96, and echocardiographic associations from 16 to 12.

Severity stratification further differentiated the cardiometabolic associations. Relative to no OSA, fully m4 adjusted ORs increased across mild, moderate, and severe OSA for essential hypertension (1.17, 1.33, and 1.58), type 2 diabetes (1.06, 1.14, and 1.25), and heart failure NOS (1.01, 1.13, and 1.48). Hypertension met Bonferroni correction in all three strata, diabetes in moderate and severe OSA, and heart failure only in severe OSA. Parallel gradients appeared in 5-year laboratory values: rank-inverse-normal β estimates for triglycerides were 0.21, 0.28, and 0.34, and those for hemoglobin A1c were 0.12, 0.20, and 0.31, respectively; all contrasts were Bonferroni-significant.

Symptoms also varied with severity, with fully adjusted continuous ESS β estimates of 0.07, 0.13, and 0.40 and ISI β estimates of 0.04, 0.06, and 0.20 across mild, moderate, and severe OSA, respectively; all met Bonferroni correction. Nevertheless, fully adjusted analyses did not show significant associations of OSA with prevalent atrial fibrillation (phecode 427.21; OR 1.04, 95% CI 0.96–1.13; P=0.300), dementia (phecode 290.1; OR 0.78, 95% CI 0.57–1.06; P=0.114), or coronary atherosclerosis (phecode 411.4; OR 1.02, 95% CI 0.96–1.08; P=0.515). Other chronic ischemic heart disease (phecode 411.8) showed a nominal positive association (OR 1.28, 95% CI 1.13–1.45; P=0.000132) that did not meet Bonferroni correction, whereas Parkinson’s disease (phecode 332) was inversely associated with OSA (OR 0.54, 95% CI 0.47–0.61; P<0.0001). Together with lower outpatient use but higher inpatient rates in the descriptive severity comparisons *(Table 2),* these findings show that increasing AHI was accompanied by selective cardiometabolic and symptom gradients rather than uniformly greater abnormality across all domains. Complete model-tier and severity estimates are provided in the *Supplementary Atlas*.

### Phase 2: incident disease and PAP use

#### Prospective associations concentrate the atlas on cardiometabolic outcomes

Prospective follow-up narrowed the broad prevalence landscape to a smaller set of predominantly cardiometabolic associations. The incidence analysis included 157,025 patients, 454,335 person-years, a median follow-up of 2.63 years, and 3,378 deaths. Among 1,013 eligible phecodes, 16 were Bonferroni-significant in m4 (11 HRs above 1 and five below 1), and 28 met the secondary FDR criterion *(Figure 8).* Beyond subsequent coding of sleep apnea itself (HR 6.36, 95% CI 6.21–6.51), elevated hazards included impaired fasting glucose (HR 1.19, 95% CI 1.15–1.24), essential hypertension (1.18, 1.14–1.22), type 2 diabetes (1.16, 1.11–1.21), hyperlipidemia (1.13, 1.09–1.18), obesity (1.13, 1.08–1.18), and heart failure (1.09, 1.05–1.13). Anxiety disorder (HR 0.89, 95% CI 0.84–0.93) and abdominal pain (0.91, 0.88–0.95) were inverse associations. Atrial fibrillation/flutter (phecode 427.2) emerged with a smaller positive association (HR 1.06, 95% CI 1.02–1.10; P=0.0041) that met neither Bonferroni nor FDR correction for the primary binary contrast. Complete results are provided in the *Supplementary Atlas (PheDAS Incidence tab)*.

**Figure 8.**
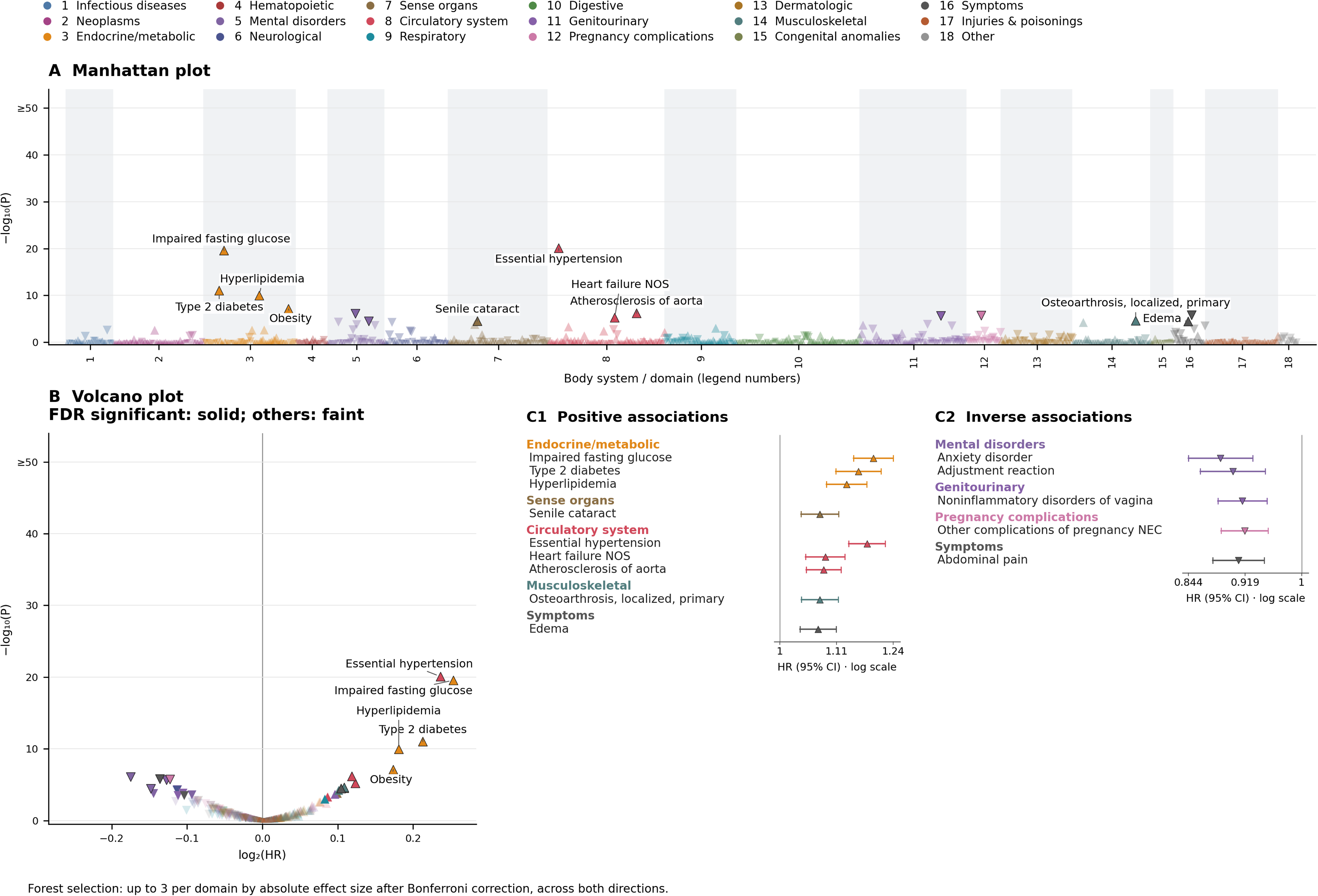
PheDAS Incidence. A, Manhattan plot; B, volcano plot; C1/C2, positive/inverse forests. PheDAS incidence analysis of 1,012 phecodes using fully adjusted (m4) AHI ≥5 versus <5 associations, excluding sleep apnea and subcodes. Complete results are provided in the **Atlas, PheDAS Incidence tab**. Colors denote body systems/domains; triangles indicate effect direction, symbol size reflects effect magnitude, and dark outlines denote Bonferroni significance. Manhattan plots label up to ten positive Bonferroni-significant associations by smallest *P* value; volcano plots label up to five by largest effect size, with solid symbols indicating FDR significance (*q*<0.05). Inverse associations are not labeled. Forest plots show up to three Bonferroni-significant features per domain, selected by absolute effect size; whiskers indicate 95% CIs. The −log10(*P*) axis is capped at 50.

#### Severity-stratified cumulative incidence

Severity gradients were apparent in absolute disease incidence, but their adjusted support differed by outcome *(Figure 9A).* Across no OSA, mild, moderate, and severe OSA, unadjusted 3-year cumulative incidence increased for essential hypertension (4.75%, 7.13%, 8.29%, 9.16%), type 2 diabetes (0.85%, 1.68%, 2.71%, 4.74%), heart failure (0.83%, 1.30%, 1.97%, 2.65%), and atrial fibrillation/flutter (0.60%, 1.00%, 1.37%, 1.69%). In fully adjusted ordinal-trend models, hypertension (HR 1.08, 95% CI 1.06–1.10), diabetes (HR 1.10, 95% CI 1.08–1.12), and heart failure (HR 1.05, 95% CI 1.03–1.07) remained Bonferroni-significant per one OSA-severity category increase (all P<0.0001). The atrial fibrillation/flutter trend was weaker (HR 1.03, 95% CI 1.01–1.05; P=0.000478) and met FDR but not Bonferroni correction.

**Figure 9.**
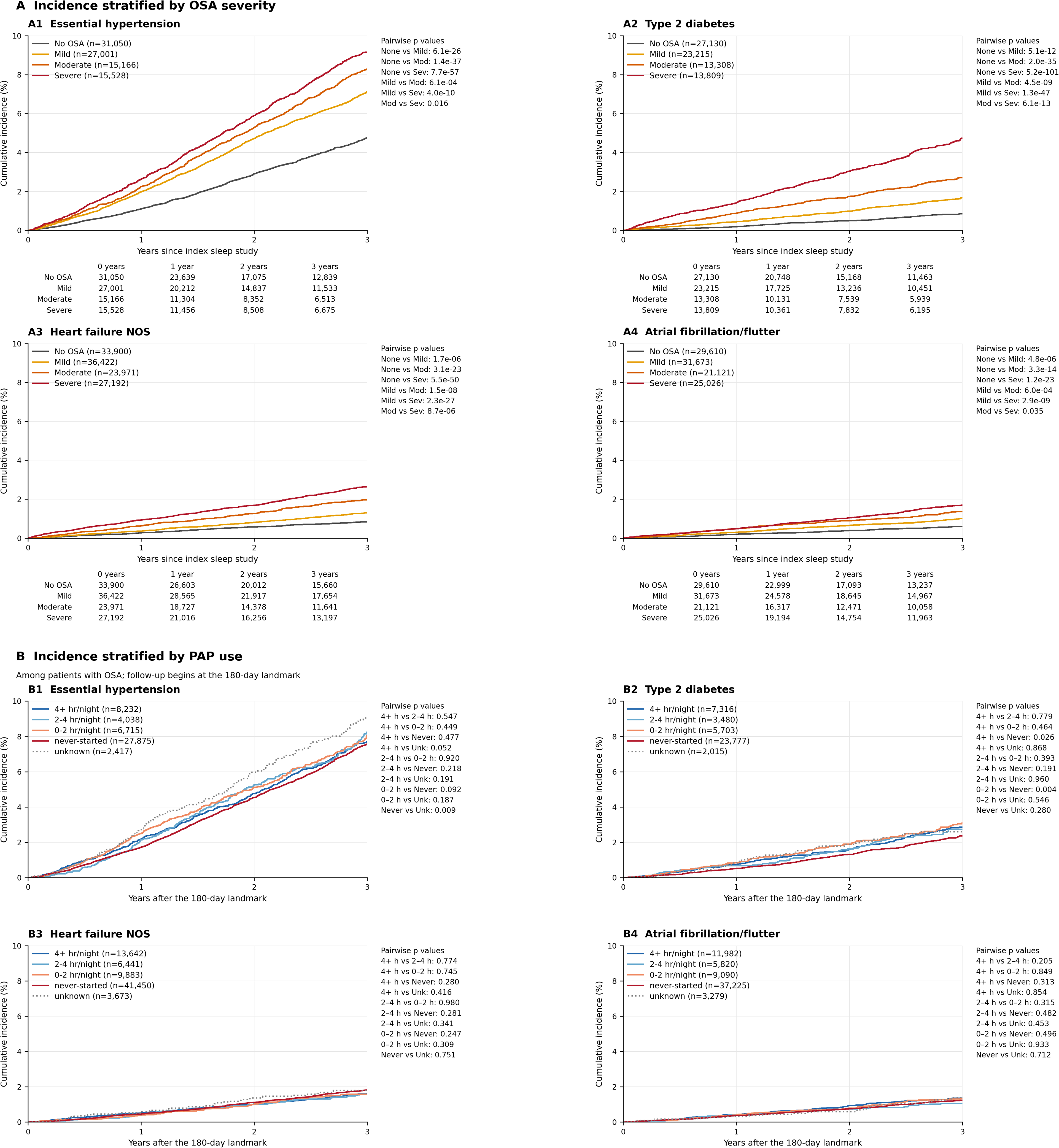
Aalen–Johansen cumulative incidence with death as a competing event. A, incidence stratified by OSA severity; A1–A4 show essential hypertension (401.1), type 2 diabetes (250.2), heart failure NOS (428.2), and atrial fibrillation/flutter (427.2), respectively. Numbers at risk at 0, 1, 2, and 3 years are shown below each severity plot. B, incidence for the same outcomes (B1–B4, respectively) after the 180-day landmark among patients with OSA, stratified by PAP-use category; unknown adherence is dotted. The PAP legends show numbers at risk at the landmark. Curves retain every incident-event step over the displayed three-year horizon. The fully adjusted PAP-use comparisons for these four outcomes were not significant after Bonferroni correction or the secondary FDR correction. Pairwise *P* values are unadjusted, two-sided cause-specific log-rank tests through 3 years, with competing deaths censored. All six OSA-severity and ten PAP-use pairwise comparisons are shown for each outcome. Pairwise-table abbreviations: None, No OSA; Mod, moderate; Sev, severe; h, hours/night; Never, never started; Unk, unknown adherence.

#### PAP-use landmark analysis

The observational PAP-use analysis did not demonstrate lower disease incidence with greater early PAP use *(Figure 9B).* In fully adjusted m4 Cox models, PAP use was not significantly associated with incident hypertension, type 2 diabetes, heart failure, or atrial fibrillation/flutter; hazard ratios across all PAP-use categories were close to 1.0 relative to ≥4 hr/night, with all 95% CIs indicating non-significance. Consistent with these adjusted findings, unadjusted 3-year Aalen–Johansen cumulative incidence after the 180-day landmark showed no clear gradient across the ≥4 hr/night, 2–3.9 hr/night, 0–1.9 hr/night, never-started, and unknown-use groups: 7.70%, 8.24%, 8.05%, 7.57%, and 9.14% for hypertension; 2.85%, 2.74%, 3.10%, 2.36%, and 2.60% for type 2 diabetes; 1.61%, 1.57%, 1.58%, 1.82%, and 1.80% for heart failure; and 1.36%, 1.05%, 1.34%, 1.24%, and 1.39% for atrial fibrillation/flutter, respectively.

### Phase 3: cross-domain phenotypes

#### Physiology symptoms and comorbidity describe different dimensions of OSA

The cross-domain analysis examined whether patients with similar AHI severity could nevertheless have different clinical profiles. Physiology, symptom, and comorbidity scores were available for 70,880 participants, who were assigned to eight strata by crossing the three median-dichotomized scores. Group sizes were well distributed, ranging from 8,349 (11.8%) for Hypoxemia Predominant to 9,249 (13.0%) for Mild All. Pairwise rank correlations between the continuous scores were close to zero: ρ=0.024 for physiology versus symptoms, 0.042 for physiology versus comorbidity, and −0.008 for symptoms versus comorbidity. Thus, the scores did not simply rank the same participants from low to high burden. The construction schematic is presented in *Supplementary Figure S1*.

The resulting profiles separated apnea frequency from coexisting disease burden *(Figure 10).* Hypoxemic Symptomatic had the highest median AHI (38.1 events/h; IQR 17.3–69.8), and 58.6% had severe OSA. By comparison, High All and Hypoxemic Comorbid each had a median of 60 distinct phecodes despite lower median AHIs of 24.9 and 21.7 events/h, respectively. *Figure 10* also provides demographic characteristics for 53,012 participants (high comorbid phenotypes were generally older in age) and follow-up descriptors for the 25,380 participants in the phenotype-incidence analysis.

**Figure 10.**
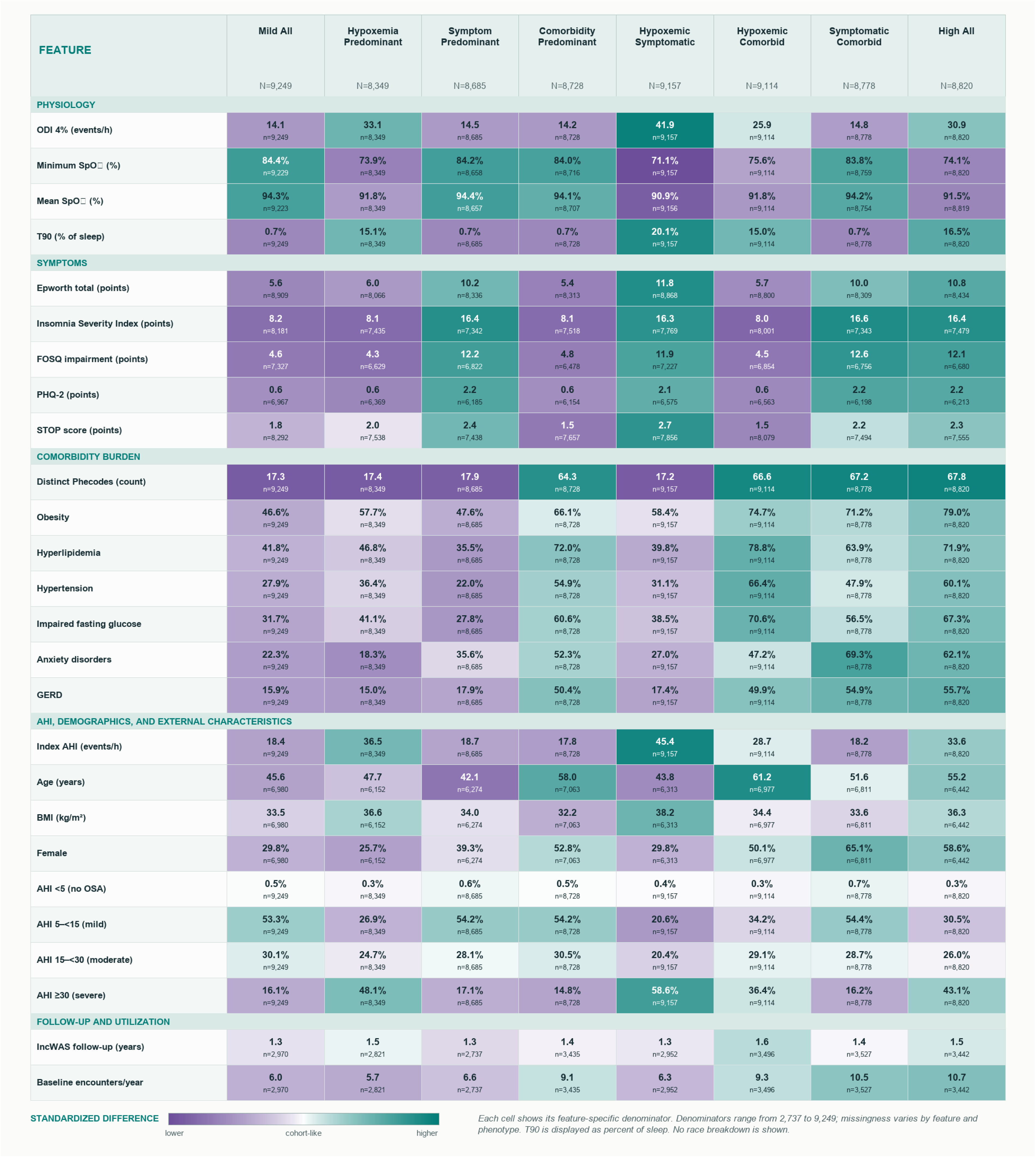
Clinical signature of the eight cross-domain phenotype octants. All 26 release-audited aggregate measures are shown across the eight octants. Cell values are native-unit means or percentages; color shows standardized difference from the shared cohort, and each cell lists its feature-specific n. Denominators are 70,880 for most phenotype measures, 53,012 for demographics, and 25,380 for follow-up and encounter measures.

#### Outcome heterogeneity extends beyond the highest AHI stratum

Subsequent disease associations differed across these clinical profiles. Of 368 eligible phecodes, 15 passed the nominal omnibus gate, yielding 120 one-versus-rest contrasts; six met Bonferroni correction and 14 met FDR correction *(Figure 11).* The significant outcomes were not confined to cardiometabolic disease. *Symptom Predominant* was associated with generalized anxiety disorder (HR 1.91, 95% CI 1.34–2.72), and *Symptomatic Comorbid* with neuralgia/neuritis/radiculitis NOS (HR 1.56, 95% CI 1.25–1.95) and gastroesophageal reflux disorder (HR 1.58, 95% CI 1.23–2.04). *High All* was associated with spondylosis (HR 1.53, 95% CI 1.18–1.98), heart failure (HR 1.58, 95% CI 1.19–2.10), and constipation (HR 1.51, 95% CI 1.16–1.97). Complete estimates are provided in the *Supplementary Atlas*.

**Figure 11.**
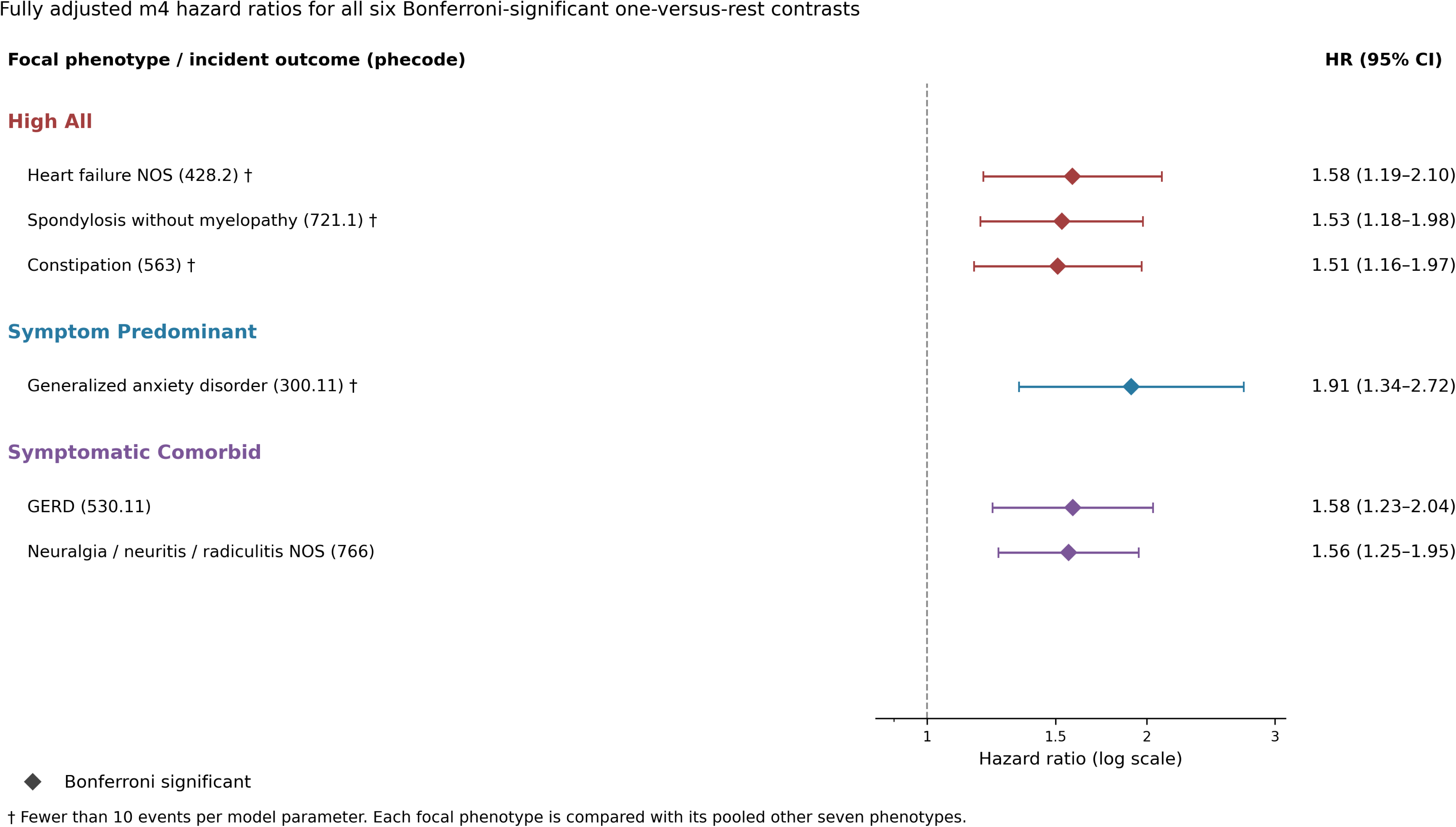
Phenotype-associated disease incidence. All six Bonferroni-significant fully adjusted (m4) cause-specific Cox contrasts are shown with 95% CIs. Of 368 eligible phecodes, 15 passed the nominal 7-df omnibus gate (*P*<0.05), yielding 120 one-versus-rest contrasts across the eight phenotypes. FDR and Bonferroni corrections were applied within each focal-phenotype family across the 15 gated outcomes. Filled diamonds denote Bonferroni significance. Fourteen contrasts met the secondary FDR criterion. All 120 contrasts are provided in the **Atlas, PheDAS Incidence tab (phenotype analyses)**.

Heart failure illustrated how multidimensional phenotype risk was not captured by median AHI alone *(Figure 12). High All* and *Hypoxemic Comorbid* had lower median AHIs (24.9 and 21.7 events/h, respectively) than *Hypoxemic Symptomatic* (38.1 events/h) yet showed higher crude 3-year heart-failure incidence. Among 18,886 participants at risk, with 164 events in the original adjusted analysis, *High All* had a higher adjusted hazard than its pooled other-seven comparator (HR 1.58, 95% CI 1.19–2.10; Bonferroni-adjusted P=0.0215), with crude cumulative incidence of 3.88% versus 1.51%. *Hypoxemic Comorbid* also had higher crude incidence (3.43% versus 1.57%), although its adjusted contrast was not significant (HR 1.18, 95% CI 0.89–1.57). In contrast, *Hypoxemic Symptomatic*, despite having the highest median AHI, had lower crude incidence (1.14% versus 1.92%) and no increased adjusted risk (HR 1.01, 95% CI 0.73–1.41). Together, these findings demonstrate that heart-failure risk across the multidimensional phenotypes did not follow the conventional ranking of AHI severity.

**Figure 12.**
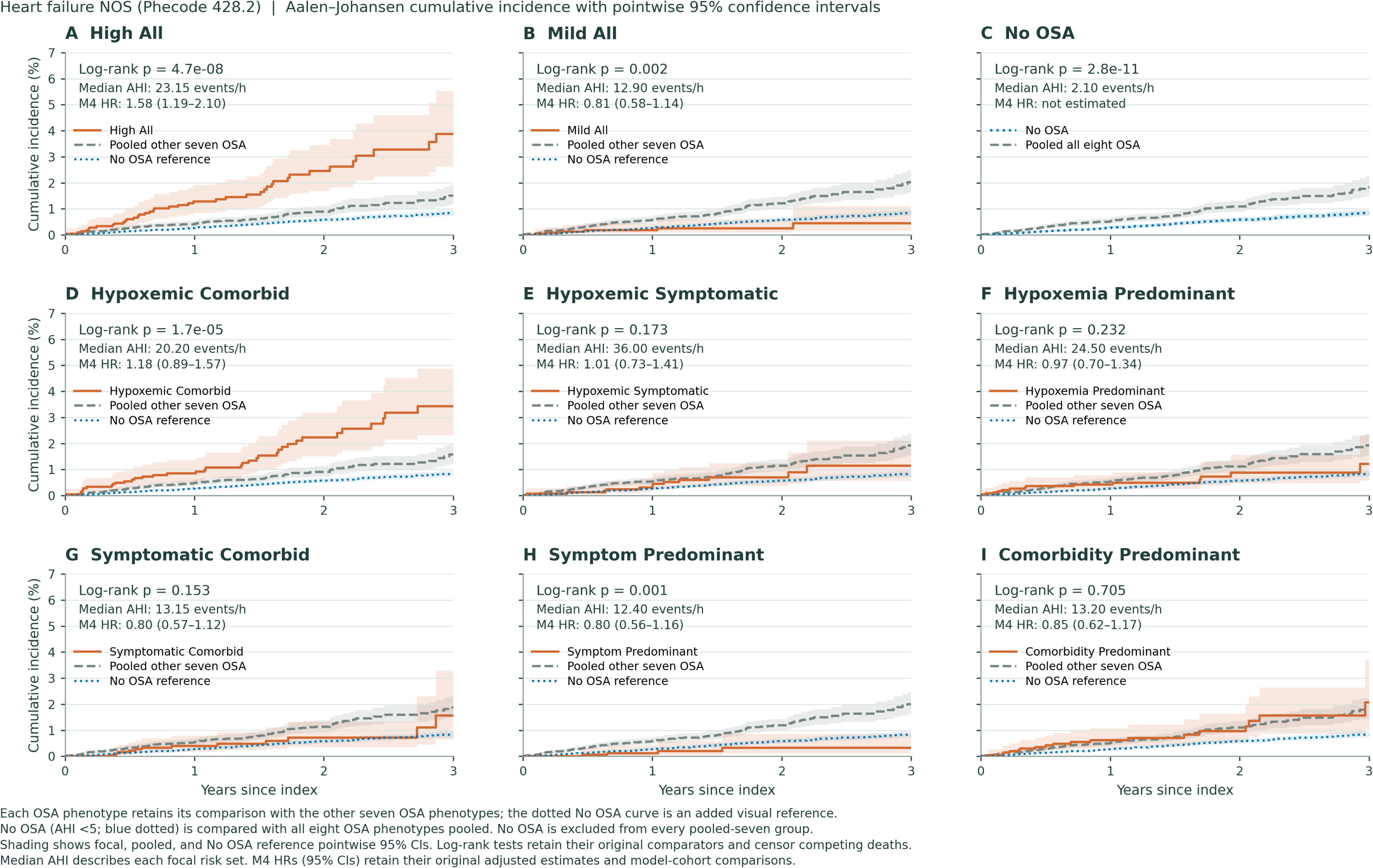
Heart failure NOS (Phecode 428.2) cumulative incidence across eight OSA phenotypes and No OSA. Each OSA phenotype is compared with the pooled other seven phenotypes; No OSA (AHI <5) is compared with all eight OSA phenotypes pooled. The No OSA curve is shown as a blue dotted reference in each phenotype panel. Curves are unadjusted Aalen–Johansen cumulative-incidence estimates with death as a competing risk; shading indicates 95% CIs. Panel *P* values are unadjusted cause-specific log-rank tests through 3 years. Median AHI describes each focal risk set. M4 HRs (95% CIs) are adjusted one-versus-rest estimates from the phenotype cohort and have <10 events per model parameter; no corresponding adjusted HR is available for No OSA versus pooled OSA.

## DISCUSSION

In this large, clinical phenome-wide association study atlas, OSA was associated with a broad but selective landscape of disease, laboratory, medication, cardiorespiratory, healthcare-utilization, behavioral, and patient-reported phenotypes. Several findings were particularly notable. First, positive associations converged most consistently across cardiometabolic domains, with higher prevalence and prospective incidence of several cardiovascular disease conditions accompanied by concordant laboratory, medication, and utilization signals. Second, the prospective incidence analysis narrowed the broad cross-sectional landscape to a more focused cardiometabolic core: hypertension, type 2 diabetes, hyperlipidemia, obesity, and heart failure. Third, 3-dimensional phenotypes constructed independently from physiologic, symptom, and comorbidity domains separated subsequent disease risk beyond traditional AHI stratification. Together, these findings support viewing OSA as a heterogeneous clinical state whose associations extend across multiple dimensions of the electronic health record.

Previous sleep phenome-wide studies have linked self-reported sleep duration, timing, and regularity^17^, wearable-derived sleep patterns^19^, and home sleep-test characteristics^18^ to diverse clinical phenotypes. More directly, Cade et al. investigated sleep apnea and disease associations using NLP-enhanced EHR phenotyping, with polysomnographic characterization in a subset.^25,26^ Our study extends this work by anchoring OSA exposure to diagnostic AHI in a substantially larger cohort and integrating diagnoses, biomarkers, treatments, healthcare utilization, symptoms, and prospective outcomes within a common analytic framework. This approach permits direct comparison of association direction, severity gradients, adjustment sensitivity, and temporality across domains.

The clearest cross-domain signal involved cardiometabolic disease. Hypertension, obesity, hyperlipidemia, impaired glucose metabolism, diabetes, and heart failure were accompanied by adverse lipid and glycemic profiles, greater use of cardiovascular and glucose-lowering medications, and echocardiographic differences in ventricular structure and function. These findings parallel those of Kohn et al., who identified associations between sleep-test characteristics and insulin resistance, lipid profiles, and cardiovascular measurements^18^. Our analysis extends this concordance to diagnoses, medication use, cardiac measurements, and prospective disease outcomes. Although consistent with established links between OSA, sympathetic activation^10,27^, intermittent hypoxemia^28–31^, and cardiometabolic dysregulation^32–34^, these observational associations do not establish causality.

The prospective analysis provides a more stringent view of this association landscape. Elevated hazards concentrated again in cardiometabolic outcomes including impaired fasting glucose, hypertension, hyperlipidemia, obesity, type 2 diabetes, and heart failure. Crude cumulative-incidence curves demonstrated dose-response relationships across conventional AHI severity categories for hypertension, diabetes, and heart failure. The smaller number of prospective incidence than prevalent associations suggests that a substantial portion of the cross-sectional phenome reflects disease already present at the time of OSA diagnosis, differences in healthcare encounter-based disease identification, or shorter duration to ascertain conditions.

Several clinically anticipated associations were notably absent or attenuated. Atrial fibrillation was null in prevalence analyses and only nominally associated with incident disease; myocardial infarction and several cerebrovascular phenotypes also did not meet significance thresholds. Incident Alzheimer’s and Parkinson’s disease were similarly null, although both models carried low-events-per-variable warnings which limit interpretation of these findings. These findings do not exclude established biologic relationships but underscore that disease-specific associations may not remain evident when evaluated under uniform adjustment, outcome definitions, and multiplicity correction, particularly for conditions with long latency to develop, low event frequency, and limited follow-up censoring duration.

More broadly, covariate adjustment, particularly for BMI, substantially attenuated the cross-sectional association landscape. Nevertheless, the cardiometabolic associations including hypertension, type 2 diabetes, and heart failure became progressively stronger across increasing OSA severity, supporting a graded relationship with disease burden. This pattern suggests that adjustment helps distinguish a more robust cardiometabolic core from a broader set of associations that may be more sensitive to demographic features, obesity, and patterns of healthcare use. The fully adjusted prevalence atlas also contained numerous inverse associations. These inverse associations should be interpreted cautiously, as they may reflect the selected nature of a sleep-tested population, referral and clinical workflow patterns, competing indications for evaluation, and diagnostic substitution rather than underlying disease biology.

Another major finding was that multidimensional phenotypes captured heterogeneity beyond AHI. Previous OSA phenotyping studies identified distinct clinical symptom presentations, including excessively sleepy, disturbed-sleep, and minimally symptomatic subtypes^2,3,35^. Polysomnographic phenotyping likewise identified cardiovascular risk profiles not captured by conventional AHI categories^5,22^. Our approach extends these studies by separately modeling physiology, symptoms, and comorbidities and integrating their nearly orthogonal burden scores into eight cross-domain phenotypes. Hypoxemic Symptomatic had the highest median AHI and greatest proportion of severe OSA, whereas High All and Hypoxemic Comorbid had lower AHIs but greater comorbidity burden. This distinction was reflected in incident heart failure: High All was associated with increased adjusted risk, whereas Hypoxemic Symptomatic was not, despite having the highest median AHI, and this contrasts previous work linking the sleepy phenotype to increased cardiovascular risk^2^. Nevertheless, our results add to the growing literature suggesting that apnea event frequency alone inadequately represents the dimensions of OSA related to symptoms^2,22,23,35^ and end-organ risk^5,22,32–34^; advanced computational analysis of raw polysomnographic signals, as exemplified by SleepFM, may eventually identify latent physiologic signatures that more directly predict subsequent clinical risk^36^. An additional nuance was that, although the symptom domain contributed independently to these multidimensional phenotypes, it did not separate into qualitatively distinct symptom subtypes. Prior clustering studies have identified patterns such as disturbed sleep versus excessive sleepiness^2,37–39^; in our cohort, symptom classes instead largely reflected increasing overall symptom burden without clear separation into distinct symptom profiles. The findings support the notion of multidimension definition of sleep quality^40,41^.

The PAP-use analysis should also be interpreted narrowly. Early PAP adherence was not significantly associated with incident hypertension, diabetes, heart failure, or atrial fibrillation. The landmark design reduced immortal-time bias by assigning adherence only after the grace period, but it could not eliminate healthy-adherer effects, treatment-selection bias, differences in therapy initiation, or residual confounding. Moreover, adherence data were available for only a subset of the cohort and were summarized from an early compliance window and does not represent treatment over years. Accordingly, these null associations should not be interpreted as evidence that PAP has no effect on long-term clinical outcomes; rather, they demonstrate the limitations of estimating treatment effects from observational adherence data even within a large integrated health system.

This study has several strengths. First, OSA exposure was anchored to objectively-measured AHI rather than diagnosis codes. Second, the cohort was large and racially and ethnically diverse, derived from an integrated healthcare setting with stable long-term membership that enabled the same patients to be characterized across an extensive, near-comprehensive health domains: diagnoses, laboratories, medications, procedures, cardiorespiratory measurements, utilization, behaviors, and symptoms. A common exposure definition, covariate ladder, and prespecified multiplicity framework allowed associations and null findings to be evaluated under comparable assumptions rather than selectively reported. Third, in addition to analyzing prevalence of outcomes, we further analyzed incident disease and incorporated analysis of potential PAP effects (although we did not discover significant effects of treatment on disease incidence). Fourth, we utilized data across domains to identify eight novel phenotypes that related to incident disease not fully captured by AHI. Finally, the release of complete aggregate summary statistics and the interactive atlas is intended to make positive as well as null findings available for external hypothesis generation.

This study has several limitations. (1) The cohort comprises patients referred for diagnostic sleep testing and is therefore not representative of the general population; the No-OSA group is a sleep-tested comparison group rather than a true healthy control population. (2) The retrospective EHR design is susceptible to residual confounding, coding error, and differential ascertainment, with potential for selection, measurement, referral, and surveillance bias. (3) Although incident outcomes required repeated post-index coding and exclusion of baseline phecode families, EHR-documented onset may not reflect true biological disease onset. (4) We did not consider apnea types such as obstructive vs central apnea. Central apnea events are relatively less common than obstructive apnea and central sleep apnea as a diagnosis was very uncommon in our cohort. Nevertheless, their endotype is different and hence may have different PheWAS profiles; for example, atrial fibrillation has been reported to be associated with central sleep apnea independent of obstructive sleep apnea^42,43^. (5) Data availability also varied across domains; for example, questionnaires were available for approximately one third of participants, while echocardiography and spirometry were available only in selected subgroups. (6) Incidence analyses were also limited by follow-up duration, an average of only 2.6 years. (7) The study was conducted within a single integrated healthcare system, and external validation is needed to determine which association and phenotype signatures generalize across populations and care settings. Nevertheless, the presented atlas provides a starting point for downstream hypothesis-driven external validation.

In conclusion, this study provides a multidomain clinical atlas of OSA, emphasizing the heterogeneous nature of OSA. AHI-defined OSA was associated with a broad clinical phenome, with the most coherent positive signals converging on cardiometabolic disease across diagnoses, laboratory biomarkers, medications, and cardiac measurements. Prospective analyses identified a more focused set of incident cardiometabolic associations, while multidimensional physiologic, symptom, and comorbidity phenotypes revealed outcome heterogeneity not captured by AHI alone. The atlas and accompanying website (https://www.osaatlas.com/) provide a resource for replication, mechanistic investigation, and hypothesis generation.

## Supporting information

Complete OSA Atlas

## Data Availability

The individual-level data underlying this study are not publicly available because they contain protected health information. All aggregate results supporting the findings of this study are provided within the manuscript and its supplementary materials.

## REFERENCES

1. Benjafield AV, Ayas NT, Eastwood PR, et al. Estimation of the global prevalence and burden of obstructive sleep apnoea: a literature-based analysis. Lancet Respir Med. Aug 2019;7(8):687–698. doi:10.1016/S2213-2600(19)30198-5

2. Mazzotti DR, Keenan BT, Lim DC, Gottlieb DJ, Kim J, Pack AI. Symptom Subtypes of Obstructive Sleep Apnea Predict Incidence of Cardiovascular Outcomes. American Journal of Respiratory and Critical Care Medicine. 2019;200(4):493–506. doi:10.1164/rccm.201808-1509OC

3. Ye L, Pien GW, Ratcliffe SJ, et al. The different clinical faces of obstructive sleep apnoea: a cluster analysis. European Respiratory Journal. 2014;44(6):1600–1607. doi:10.1183/09031936.00032314

4. Zinchuk AV, Gentry MJ, Concato J, Yaggi HK. Phenotypes in obstructive sleep apnea: A definition, examples and evolution of approaches. Sleep Medicine Reviews. 2017;35:113–123. doi:10.1016/j.smrv.2016.10.002

5. Zinchuk AV, Jeon S, Koo BB, et al. Polysomnographic phenotypes and their cardiovascular implications in obstructive sleep apnoea. Thorax. 2018;73(5):472–480. doi:10.1136/thoraxjnl-2017-210431

6. Edwards C, Almeida OP, Ford AH. Obstructive sleep apnea and depression: A systematic review and meta-analysis. Maturitas. 2020;142:45–54. doi:10.1016/j.maturitas.2020.06.002

7. Hou H, Zhao Y, Yu W, et al. Association of obstructive sleep apnea with hypertension: A systematic review and meta-analysis. Journal of Global Health. 2018;8(1):010405. doi:10.7189/jogh.08.010405

8. Reutrakul S, Mokhlesi B. Obstructive Sleep Apnea and Diabetes: A State of the Art Review. Chest. 2017;152(5):1070–1086. doi:10.1016/j.chest.2017.05.009

9. Wu Z, Chen F, Yu F, Wang Y, Guo Z. A meta-analysis of obstructive sleep apnea in patients with cerebrovascular disease. Sleep and Breathing. 2018;22(3):729–742. doi:10.1007/s11325-017-1604-4

10. Yeghiazarians Y, Jneid H, Tietjens JR, et al. Obstructive Sleep Apnea and Cardiovascular Disease: A Scientific Statement From the American Heart Association. Circulation. 2021;144(3):e56–e67. doi:10.1161/CIR.0000000000000988 PMID - 34148375

11. Zhang D, Ma Y, Xu J, Yi F. Association between obstructive sleep apnea (OSA) and atrial fibrillation (AF): A dose-response meta-analysis. Medicine. 2022;101(30):e29443. doi:10.1097/MD.0000000000029443

12. Denny JC, Bastarache L, Ritchie MD, et al. Systematic comparison of phenome-wide association study of electronic medical record data and genome-wide association study data. Nature Biotechnology. 2013;31(12):1102–1110. doi:10.1038/nbt.2749

13. Denny JC, Ritchie MD, Basford MA, et al. PheWAS: demonstrating the feasibility of a phenome-wide scan to discover gene-disease associations. Bioinformatics. 2010;26(9):1205–1210. doi:10.1093/bioinformatics/btq126

14. Denny JC, Bastarache L, Roden DM. Phenome-Wide Association Studies as a Tool to Advance Precision Medicine. Annual Review of Genomics and Human Genetics. 2016;17:353–373. doi:10.1146/annurev-genom-090314-024956

15. Chaganti S, Welty VF, Taylor W, et al. Discovering novel disease comorbidities using electronic medical records. PLOS ONE. 2019;14(11):e0225495. doi:10.1371/journal.pone.0225495

16. Kerley CI, Chaganti S, Nguyen TQ, et al. pyPheWAS: A Phenome-Disease Association Tool for Electronic Medical Record Analysis. Neuroinformatics. 2022;20(2):483–505. doi:10.1007/s12021-021-09553-4

17. Dashti HS, Cade BE, Stutaite G, Saxena R, Redline S, Karlson EW. Sleep health, diseases, and pain syndromes: findings from an electronic health record biobank. Sleep. 2021;44(3):zsaa189. doi:10.1093/sleep/zsaa189 PMID - 32954408 PMCID - PMC7953212

18. Kohn S, Diament A, Godneva A, et al. Phenome-wide associations of sleep characteristics in the Human Phenotype Project. Nature Medicine. 2025;31(3):1026–1037. doi:10.1038/s41591-024-03481-x PMID - 39870817

19. Zheng NS, Annis J, Master H, et al. Sleep patterns and risk of chronic disease as measured by long-term monitoring with commercial wearable devices in the All of Us Research Program. Nature Medicine. 2024;30(9):2648–2656. doi:10.1038/s41591-024-03155-8 PMID - 39030265 PMCID - PMC11405268

20. Dennis JK, Sealock JM, Straub P, et al. Clinical laboratory test-wide association scan of polygenic scores identifies biomarkers of complex disease. Genome Medicine. 2021;13(1):6. doi:10.1186/s13073-020-00820-8

21. Ryan PB, Madigan D, Stang PE, Schuemie MJ, Hripcsak G. Medication-wide association studies. CPT: Pharmacometrics & Systems Pharmacology. 2013;2(9):e76. doi:10.1038/psp.2013.52

22. Malhotra A, Ayappa I, Ayas N, et al. Metrics of sleep apnea severity: beyond the apnea-hypopnea index. Sleep. 2021;44(7):zsab030. doi:10.1093/sleep/zsab030

23. Zinchuk AV, Yaggi HK. Phenotypic Subtypes of OSA: A Challenge and Opportunity for Precision Medicine. Chest. 2020;157(2):403–420. doi:10.1016/j.chest.2019.09.002

24. Wei W-Q, Bastarache LA, Carroll RJ, et al. Evaluating phecodes, clinical classification software, and ICD-9-CM codes for phenome-wide association studies in the electronic health record. PLOS ONE. 2017;12(7):e0175508. doi:10.1371/journal.pone.0175508 PMID - 28686612 PMCID - PMC5501393

25. Cade BE, Hassan SM, Dashti HS, et al. Sleep apnea phenotyping and relationship to disease in a large clinical biobank. JAMIA Open. Apr 2022;5(1):ooab117. doi:10.1093/jamiaopen/ooab117

26. Cade BE, Li L, Duff M, et al. Associations between obstructive sleep apnea and comorbidities in a large clinical biobank. medRxiv. Aug 1 2025;doi:10.1101/2025.07.30.25332466

27. Dissanayake HU, Bin YS, Ucak S, de Chazal P, Sutherland K, Cistulli PA. Association between autonomic function and obstructive sleep apnea: A systematic review. Sleep Medicine Reviews. 2021;57:101470. doi:10.1016/j.smrv.2021.101470 PMID - 33839505

28. Gilmartin GS, Lynch M, Tamisier R, Weiss JW. Chronic intermittent hypoxia in humans during 28 nights results in blood pressure elevation and increased muscle sympathetic nerve activity. American Journal of Physiology-Heart and Circulatory Physiology. 2010;299(3):H925–H931. doi:10.1152/ajpheart.00253.2009 PMID - 20581089

29. Leuenberger UA, Brubaker D, Quraishi SA, Hogeman CS, Imadojemu VA, Gray KS. Effects of intermittent hypoxia on sympathetic activity and blood pressure in humans. Autonomic Neuroscience. 2005;121(1-2):87–93. doi:10.1016/j.autneu.2005.06.003 PMID - 15996901

30. Leuenberger UA, Hogeman CS, Quraishi SA, Linton-Frazier L, Gray KS. Short-term intermittent hypoxia enhances sympathetic responses to continuous hypoxia in humans. Journal of Applied Physiology. 2007;103(3):835–842. doi:10.1152/japplphysiol.00036.2007 PMID - 17556498

31. Prabhakar NR, Peng Y-J, Nanduri J. Carotid body hypersensitivity in intermittent hypoxia and obstructive sleep apnoea. The Journal of Physiology. 2023;601(24):5481–5494. doi:10.1113/JP284111 PMID - 37029496

32. Azarbarzin A, Sands SA, Stone KL, et al. The hypoxic burden of sleep apnoea predicts cardiovascular disease-related mortality: the Osteoporotic Fractures in Men Study and the Sleep Heart Health Study. European Heart Journal. 2019;40(14):1149–1157. doi:10.1093/eurheartj/ehy624 PMID - 30376054

33. Azarbarzin A, Sands SA, Taranto-Montemurro L, et al. The Sleep Apnea-Specific Hypoxic Burden Predicts Incident Heart Failure. Chest. 2020;158(2):739–750. doi:10.1016/j.chest.2020.03.053 PMID - 32298733

34. Redline S, Azarbarzin A, Peker Y. Obstructive sleep apnoea heterogeneity and cardiovascular disease. Nature Reviews Cardiology. 2023;20(8):560–573. doi:10.1038/s41569-023-00846-6 PMID - 36899115

35. Keenan BT, Kim J, Singh B, et al. Recognizable clinical subtypes of obstructive sleep apnea across international sleep centers: a cluster analysis. Sleep. Mar 1 2018;41(3)doi:10.1093/sleep/zsx214

36. Thapa R, Kjaer MR, He B, et al. A multimodal sleep foundation model for disease prediction. Nature Medicine. 2026;32(2):752–762. doi:10.1038/s41591-025-04133-4 PMID - 41495409

37. Keenan BT, Kim J, Singh B, et al. Recognizable clinical subtypes of obstructive sleep apnea across international sleep centers: a cluster analysis. Sleep. 2018;41(3):zsx214. doi:10.1093/sleep/zsx214 PMID - 29315434

38. Keenan BT, Ye L, Pien GW, et al. Symptom subtypes of obstructive sleep apnea 10 years later: past, present, and future. Sleep. 2025;48(7):zsaf082. doi:10.1093/sleep/zsaf082 PMID - 40202500

39. Ye L, Pien GW, Ratcliffe SJ, et al. The different clinical faces of obstructive sleep apnoea: a cluster analysis. European Respiratory Journal. 2014;44(6):1600–1607. doi:10.1183/09031936.00032314 PMID - 25186268

40. Buysse DJ. Sleep Health: Can We Define It? Does It Matter? Sleep. 2014;37(1):9–17. doi:10.5665/sleep.3298 PMID - 24470692 PMCID - PMC3902880

41. Knutson KL, Phelan J, Paskow MJ, et al. The National Sleep Foundation’s Sleep Health Index. Sleep Health. 2017;3(4):234–240. doi:10.1016/j.sleh.2017.05.011 PMID - 28709508

42. Harmon EK, Stafford P, Ibrahim S, et al. Atrial fibrillation is associated with central sleep apnea in clinic patients undergoing diagnostic polysomnography. Journal of Arrhythmia. 2020;36(6):991–996. doi:10.1002/joa3.12427 PMID - 33335615 PMCID - PMC7733563

43. Tung P, Levitzky YS, Wang R, et al. Obstructive and Central Sleep Apnea and the Risk of Incident Atrial Fibrillation in a Community Cohort of Men and Women. Journal of the American Heart Association. 2017;6(7):e004500. doi:10.1161/JAHA.116.004500 PMID - 28668820 PMCID - PMC5586257

