## Supplementals for "A Clinical Atlas for Phenome-Wide Association Study of Obstructive Sleep Apnea Across Electronic Health Record Domains"

### Supplementary Material

Complete association results are provided separately in the **Supplementary Atlas** Excel workbook. Its tabs are PheDAS Prevalence, PheDAS Incidence, LabWAS, MedWAS, ProcWAS, CRMWAS, UtilWAS, BehWAS, and QWAS.

#### Supplementary Methods

##### Study design and setting

We conducted a retrospective cohort study of adults who underwent diagnostic sleep testing within Kaiser Permanente Southern California (KP), an integrated delivery system whose EHR (Epic Systems, Verona, Wisconsin) captures encounters, diagnoses, laboratory results, medication dispensing, procedures, cardiopulmonary measurements and questionnaires for a stable insured membership. The study was conducted as part of the Human Sleep Project (HSP) under protocols approved by the BIDMC Institutional Review Board (#2024P000804/#2022P000417), which granted a waiver of informed consent for this retrospective study. Reporting follows the STROBE framework.

##### Cohort and index date

Adults (≥18 years) were included for analysis if they had available diagnostic sleep study data from polysomnography (PSG) or home sleep apnea testing (HSAT) for studies dated 2016–2025. The earliest sleep study date (if multiple) was selected as the index date and anchors every exposure, covariate, and outcome window. Patients were excluded if they were missing any demographic information (i.e., race/ethnicity), body mass index (BMI), fewer than two pre-index date diagnostic encounters (to ensure minimal opportunity for proper comorbidity coding), any encounter coded with OSA ICD (ICD-10 G47.3x or ICD-9 327.2x) before index.

##### Exposure

OSA status and severity were based on the index apnea-hypopnea index (AHI) utilizing 4% hypopnea criteria, utilizing standard thresholds: none (<5; reference), mild (5 to <15), moderate (15 to <30), severe (≥30). All analyses use the same contrasts: omnibus (joint Wald test), ordinal trend, three pairwise contrasts against none (i.e., Mild vs None; Moderate vs None; Severe vs None), AHI ≥5 vs <5 (OSA vs No OSA), and AHI ≥15 vs <5 (Moderate-Severe vs None OSA).

##### Covariates and the adjustment ladder

We use a nested ladder for covariate adjustments: m1 is unadjusted; m2 is adjusted for age, sex, race/ethnicity, index year; m3 is adjusted for m2 covariates plus smoking status, median household income group by region; m4 is adjusted for m3 covariates plus BMI. Smoking and income retain "unknown" and "missing" as explicit modeled levels (i.e., no data was imputed). Continuous covariates were z-standardized on the full cohort before any per-feature subsetting. Due to similar findings between m2 and m3 adjustments, LabWAS, MedWAS, UtilWAS, BehWAS report m1/m3/m4; the remainder report m1–m4. The adjustment ladder and severity contrasts are provided in the **Supplementary Atlas (PheDAS Prevalence tab)**.

##### Common analytic framework

Each domain converts the record into per-patient features on a defined pre-index window, then regresses each feature on the exposure across the ladder. Continuous features are rank-inverse-normal transformed and fitted by ordinary least squares, so every β is in standard-deviation units and comparable across features of different scale; binary features by logistic regression (odds ratio); rate features by Poisson regression with offset log observation-years and HC0 robust standard errors (incidence rate ratio). The default inclusion floor is ≥500 patients per feature, with ≥20 per severity stratum required for any contrast.

Benjamini–Hochberg false-discovery-rate (FDR) control and Bonferroni correction were applied at α = 0.05 within each family, a family being one (domain × window × contrast × covariate model × effect type). Families are never pooled: domains differ in denominator, in the meaning of a null, and in the number of features tested, so a single global correction would not be interpretable.

#### Phase 1 – PheDAS prevalence

##### Phase 1a — Disease domain association study

All ICD-9-CM and ICD-10-CM diagnoses recorded on or before index were extracted, giving 27.7 million distinct patient × code records, and mapped to phecodes with the pyPheWAS phecode map v1.2 (ICD-9 plus ICD-10 v1.2-beta). A patient is a case for a phecode if they carry ≥1 mapped code before index. In total, 1,802 phecodes were tested. Each phecode was regressed on the exposure by maximum likelihood estimation (MLE) logistic regression, the default used for the rest of the study.

##### Phase 1b — Multi-domain association sweeps

Phase 1b domains use the same cohort spine, index date, exposure contrasts, covariate ladder and estimators as Phase 1a. Unless specified otherwise, features were built on two pre-index windows: index minus 1 year (1-year window) and index minus 5 years (5-year window).

##### LabWAS (Laboratory Values)

2,079 free-text analyte names were mapped to LOINC 2.82 by an automated matcher with verified clinical curation, and units were derived from value distributions and corrected using explicit conversion rules (**Supplementary Methods**). LabWAS consists of three outcomes: (i) values, the per-patient median among tested patients only; (ii) order rate, count per observation-year among tested patients, by Poisson regression; and (iii) ordering propensity, any order versus none over the full cohort, by MLE logistic regression. Constructs ii and iii are components of a two-part ordering hurdle decomposition and analyze utilization rather than physiology.

##### MedWAS (Medications)

Outpatient pharmacy records were collapsed to distinct fills and grouped at the four-digit Generic Product Identifier (GPI) drug class. A patient is 1 if they filled any drug in that class during the window and 0 otherwise.

#### ProcWAS (Procedures)

Procedures were mapped from primary Current Procedural Terminology (CPT) codes and a large language model (Claude Opus 4.8) created a curated crosswalk with a descriptor, procedure type, and body system. The crosswalk was manually reviewed for accuracy by a physician (DH). Procedures regarding sleep medicine and OSA treatment (e.g. polysomnography, home sleep apnea test, etc.) were explicitly excluded. The outcome was procedures per year by Poisson regression.

#### CRMWAS (Cardiorespiratory Measurements)

Two measurement streams sit outside the coded vocabularies and were analysed on the 5-year window as a distinct arm: (1) echocardiography (22 parameters of chamber size, mass, systolic and diastolic function, aortic dimension and estimated pulmonary pressure) and (2) spirometry (14 pre- and post-bronchodilator parameters). Unlike the coded domains, this arm is complete-case and not zero-filled (i.e., a patient without an echocardiogram has an “unknown” ejection fraction rather than “zero”) — so analysed n varies by feature and is reported on every row (echocardiography 3,577–46,223; spirometry 5,152–7,874). Per-patient values are the mean of in-window studies after unit repair and gating, fitted across m1 through m4 with index year mandatory, and multiplicity was corrected within each arm separately. Resting systolic blood pressure was treated as a positive control and kept out of the covariate set for other features. The spirometry arm is exploratory: only 3.6% of the cohort contributed a measurement, the source terminates in March 2019 while index dates run to March 2025, and coverage is confounded with index year. Selection into the measured sample is not severity-neutral in either arm; therefore, no estimate here is population-marginal.

##### Healthcare utilization (UtilWAS)

Encounters were defined as distinct utilization identifiers and classified into seven care categories: physician-level outpatient, allied-health outpatient, emergency department, inpatient, urgent care, secure message/telephone, and other. Two families were fitted: (1) specialty encounter rates, encounters per year within each department specialty over the full cohort (102 specialties at 5 years, 80 at 1 year) and (2) phecode utilization, in which for each of seven care-setting × diagnosis-position combinations (physician primary; emergency, urgent-care and inpatient, each primary and secondary) a two-part model separates presence of the phecode in that setting from its count among carriers, which must not be combined into one effect. Features required ≥500 patients, giving 38,303 tests at 5 years and 11,468 at 1 year.

##### Health behaviours (BehWAS)

Five outcomes were built, each in binary and continuous form: (1) medication adherence (proportion of days covered [PDC] as a continuous variable, and binary adherence defined as PDC ≥0.80; both analysed among anti-hypertensive and anti-cholesterol medication users, as PDC is undefined without exposure to therapy; n=96,086) ; (2) influenza vaccination (n=138,443, 62.3%); (3) medical-leave claims under the Family and Medical Leave Act (filed, adjudicated, pregnancy-related, duration; 2,970 at 1 year, 10,885 at 5 years); (4) appointment non-attendance (13,843, 6.2%); (5) days off- work (5,962, 2.7%). The first three were windowed (1-year and 5 years); the last two carry no usable event date and are lifetime per-person values that also incorporate post-index experience, an asymmetry with every other Phase 1b feature and a reason to read them as descriptive. All outcomes except adherence are defined over the whole cohort with non-events zero. Correction within this domain is reported for completeness rather than used as a filter; the domain is designated preliminary in the release.

##### Patient-reported symptoms (QWAS)

Patient-reported data were collected via sleep questionnaires (“Sleep Intake Questionnaire”) from three sources: (1) the EHR (2) a sleep-laboratory management system (Somnoware, Resmed Corp, San Diego, CA); (3) scanned intake forms. The data were harmonized into one encounter-grain table (228,790 encounters, 158,324 patients; **Supplementary Methods**). One questionnaire per patient was selected as the encounter closest to and on or before index; post-index questionnaires were excluded, as they may follow positive-airway-pressure initiation and would induce reverse causation. The analytic sample is 74,061 patients, 33.3% of the cohort. Unlike a phecode, an unanswered item is unknown rather than a structural zero, so this domain does not share the whole-cohort denominator: analysis is complete-case per item with n reported on every row (26,372–73,102). After floors of ≥500 answered patients and ≥100 in the minority class of a binary feature, 92 features were retained across ten content domains– individual items plus derived scores for the Epworth Sleepiness Scale (ESS), Insomnia Severity Index (ISI), Patient Health Questionnaire-2 (PHQ-2) and Functional Outcomes of Sleep Questionnaire 10 item (FOSQ-10). Each feature was tested in both applicable forms, the binary form cut at a uniform "moderate or worse" scale midpoint for ordinal items and at published cut-points for composites (ESS ≥11, ISI ≥15, PHQ-2 ≥3). Because 40 items contribute both forms, the odds-ratio phenome (83 features) and the β phenome (49 features) are corrected as separate families, giving 4,620 tests. Items that themselves drive referral for sleep testing (snoring, witnessed apnea, gasping) were retained but flagged as ascertainment-sensitive.

#### Phase 2 — PheDAS incidence

Phase 2 analyzes whether baseline OSA severity predicts new onset of each phecode among patients free of it at index (utilizing the same spine, exposure and adjustment ladder m1-m4).

##### Event store and incident-case definition

A dated phecode event store was built in a single streaming pass over the utilization table, restricted to service dates on or before the administrative cutoff of 31 May 2023, the last date for which a full-phenome diagnosis extract exists. Codes were mapped with the same v1.2 maps as Phase 1, with one correction: because the ICD-10-beta map often lists three-character stems where billing uses fully specified codes, codes were matched exactly where possible and otherwise by longest map prefix, recovering approximately 29,000 codes that exact matching silently dropped. An incident event is the date of the second distinct post-index occurrence of a phecode, a rule of two that suppresses transient rule-out coding; codes on the index day count as baseline. Because the v1.2 ICD-9 and ICD-10 maps assign the same disease to different phecodes by coding era, a leaf-level washout would misclassify ICD-9-era prevalent disease as incident (measured leakage up to 37%); the at-risk set therefore excludes patients with a baseline occurrence of that phecode or of any phecode in its standard control-exclusion family.

##### Follow-up and censoring

Follow-up begins at index on a time-since-index scale, censored at the earliest of the administrative cutoff, the end of insurance coverage where it binds before that cutoff (25,904 patients, 16%, disenrolled early and are censored rather than miscounted as event-free), and death, treated as a competing event with all models cause-specific. Because the outcome extract ends before the last index dates, 65,084 patients contribute no observable follow-up; this censoring is calendar-driven and therefore non-informative. The incidence spine is 157,025 patients contributing 454,335 person-years (median follow-up 2.63 years) and 3,378 deaths (2.2%).

##### Estimation

Each phecode was fitted by cause-specific Cox proportional-hazards regression with a small ridge penalty (0.1) for stability on rare outcomes, across m1–m4. Here m3 additionally includes the log baseline encounter rate, a surveillance-bias control: more severe OSA generates more clinical contact and therefore more opportunity for a diagnosis to be recorded. Phecodes with ≥20 incident events were retained, giving 1,013 phecodes and 28,364 model fits; fits with fewer than ten events per parameter, or that failed to converge, are flagged. Proportional hazards were assessed by Schoenfeld residual tests and reported as a flag rather than a gate, since at this sample size the test detects departures of no practical consequence. Positive-airway-pressure treatment is not adjusted for, by design: adjusting for this would block the potential causal pathway of interest, as it acts as a possible mediator between OSA and the outcome.

##### Absolute risk

Because hazard ratios do not convey burden, crude incidence densities per 1,000 person-years were computed by severity band and by year-interval since index (0–1, 1–2, 2–3 years), Cumulative incidence functions were estimated using the Aalen–Johansen estimator in the presence of competing risks, as the complement of Kaplan–Meier survival curves is known to overestimate absolute risk in this setting.

##### Positive-airway-pressure adherence

Adherence derives from a therapy-anchored compliance summary covering the first 90 days of therapy, available for 38.5% of the cohort; no per-night usage series exists, which constrains the design. A naive adherent-versus-non-adherent comparison is subject to immortal-time bias (a patient must survive and accrue usage to be labelled adherent) and to healthy-adherer confounding. We therefore used a grace-period landmark design: follow-up begins at index plus a grace period, onsets within that window are reclassified prevalent, only patients alive, event-free and enrolled at the landmark enter, and adherence is a fixed label assigned at the landmark using only information available within the grace window. Because therapy begins a median of 20 days after the diagnostic study, 180 days is the primary grace period; at 90 days the compliance window would overhang the landmark for most treated patients and the usage levels degenerate. Usage levels (≥4 h, 2–4 h, <2 h per night) were assigned only where the compliance window fell entirely within the grace window; other patients were coded unknown or never-started. Models were restricted to patients with OSA, adjusted for m3/m4 plus ordinal severity, and fitted as cause-specific Cox models for incident phecodes and all-cause mortality, alongside clinically unrelated negative-control outcomes estimating the residual healthy-adherer confounding floor against which any apparent adherence effect must be read. The landmark design removes immortal time; it does not make the comparison causal.

#### Phase 3 — Cross-domain phenotype construction and incidence

##### Domain-specific latent models

Three domains were modeled independently. The physiologic model used 10 standardized sleep-study indicators spanning oxygen desaturation, saturation burden, pulse-rate measures, and positional physiology (maximal N=239,248) and used full-information maximum likelihood under a missing-at-random assumption. The symptom model used 40 questionnaire indicators covering sleepiness, insomnia, function, mood screening, and related sleep complaints (maximal N=74,433). The comorbidity model used 172 baseline chronic-disease phecode indicators (maximal N=235,861) after rolling leaf codes to logical-OR parent features, excluding OSA/sleep codes, nonchronic categories, administrative/process codes, and sex-specific anatomy, and requiring prevalence ≥2%. The comorbidity domain is a coded-burden gradient and may reflect healthcare contact as well as disease.

For each domain, one- through eight-class solutions were screened using a predeclared rule: the BIC elbow was computed among nondegenerate fits; relative entropy had to be ≥0.70; the smallest class had to include ≥1% and ≥500 participants; the best likelihood had to replicate across independent starts; no standardized component variance could be <0.01; and split-half adjusted Rand index had to be ≥0.70. Forty short starts on a 25,000-person subsample ranked initializations, after which the six best starts were run to convergence in the full domain cohort. A three-class solution was selected for each domain (relative entropy 0.925, 0.792, and 0.917; split-half adjusted Rand index 0.966, 0.926, and 0.981 for physiology, symptoms, and comorbidity). AHI was not an indicator; it was related to the fitted classes only as an external descriptor using a three-step BCH correction.

##### Cross-linking into eight phenotype octants

Posterior probabilities were converted to posterior-weighted ordered domain scores, with higher values indicating greater physiologic, symptom, or comorbidity burden. The 70,880 participants scored in all three domains formed the shared phenotype cohort. Each score was split at its shared-cohort median (physiology 1.0022558, symptoms 1.0041043, comorbidity 0.9958656), producing eight 2×2×2 octants with complete classification. The cuts are cohort-specific descriptive boundaries, not transferable clinical thresholds. Octants were named Mild All, Hypoxemia Predominant, Symptom Predominant, Comorbidity Predominant, Hypoxemic Symptomatic, Hypoxemic Comorbid, Symptomatic Comorbid, and High All. Pairwise continuous-score Spearman correlations were 0.024, 0.042, and −0.008; modal class-label Cramér's V ranged from 0.011 to 0.032. **Supplementary Figure S1** shows construction, and **Figure 10** shows release-audited aggregate characteristics. Demographics cover 53,012 complete study-spine participants, and incidence follow-up/encounter descriptors cover the 25,380-person incidence spine; other metrics use their available aggregate denominators.

##### Phenotype-incidence models

The phenotype-incidence spine included 25,380 participants, 35,994 person-years, and mean follow-up of 1.42 years. Outcomes used the Phase 2 rule-of-two incident date and control-exclusion-family baseline washout; phecodes 327.x and 347.x were excluded as circular with the sleep-study cohort. The m4 cause-specific Cox model included an age spline, sex, centered index year, race/ethnicity, smoking, neighborhood income, baseline observation-window length, log baseline encounter rate, and BMI. Death was a competing event. Octant models used ridge penalization 0.01, with fallback to 0.1 if required; the penalizer and events-per-variable warning were retained with every result. PAP use was not adjusted because it may lie on the pathway between phenotype and outcome.

Testing was hierarchical. Stage 1 fit one eight-level exposure model per outcome and used a 7-df joint Wald test. Outcomes with raw omnibus $P<0.05$ entered Stage 2, which fit each octant against the pooled other seven. Benjamini–Hochberg q values and Bonferroni flags were calculated separately within each focal-octant contrast across the 15 gated phecodes; therefore, Bonferroni was not 0.05 divided by all 120 contrasts. Of 368 phecodes tested at m4, 15 entered Stage 2, yielding the complete 120-row gated set in the **Supplementary Atlas (PheDAS Incidence tab, phenotype analyses)**. One omnibus outcome, neuralgia/neuritis/radiculitis NOS, was FDR-significant; 14 Stage-2 contrasts were FDR-significant and six met Bonferroni correction. **Figure 11** displays all 14 FDR-significant contrasts with 95% CIs and Bonferroni and low-events-per-parameter indicators.

##### Absolute-risk curves and interpretation

Selected curves are unadjusted, tie-aware Aalen–Johansen cumulative-incidence functions on a 0–3-year grid with death competing; censoring at an event time remains in that time's risk set. The annotated hazard ratios are adjusted m4 estimates and must not be interpreted as curve-derived or pairwise effects. Each panel compares one focal phenotype with its own pooled other-seven group. Mean follow-up was short and only approximately 8.7% of pooled baseline risk sets remained observed at 3 years; risk tables and the EPV<10 flag should therefore accompany endpoint values. Proportional-hazards diagnostics were not computed for octant models, and the observational associations do not establish biological subtypes or causal effects.

##### Software and reproducibility

Analyses used Python 3.12.7 with DuckDB 1.2.1 for out-of-core extraction from the multi-terabyte source exports, statsmodels 0.14.5 for generalised linear and least-squares models, lifelines for Cox regression and Aalen–Johansen estimation, scipy 1.16.0 for the non-parametric screens, and pyPheWAS 4.2.0, which supplies both the ICD PheDAS regression engine and the phecode map v1.2 (ICD-10 v1.2-beta) and control-exclusion resources used by Phase 2; numpy 2.2.6, pandas 2.3.3. Laboratory harmonisation used LOINC release 2.82 (integrity hash recorded with the crosswalk). Code is organised as ordered, re-runnable stages per domain with all thresholds, windows and covariate specifications as module-level constants, and is available at [Repository URL/DOI to be added].

##### Data availability

Complete summary statistics for every domain (effect estimate, standard error, confidence interval, P value, per-family FDR and Bonferroni flags, and analysed n) are provided in the separate **Supplementary Atlas** Excel workbook and through an interactive browser at OSAatlas.com, archived with a persistent identifier at [DOI to be added]. The release is aggregate only: exposure-by-outcome cells are suppressed under an institutionally approved disclosure protocol in which counts below 11 are returned as null rather than zero, complementary cells are suppressed jointly to prevent reconstruction by differencing, and cumulative event totals are withheld for the survival products.

### Phenotype construction

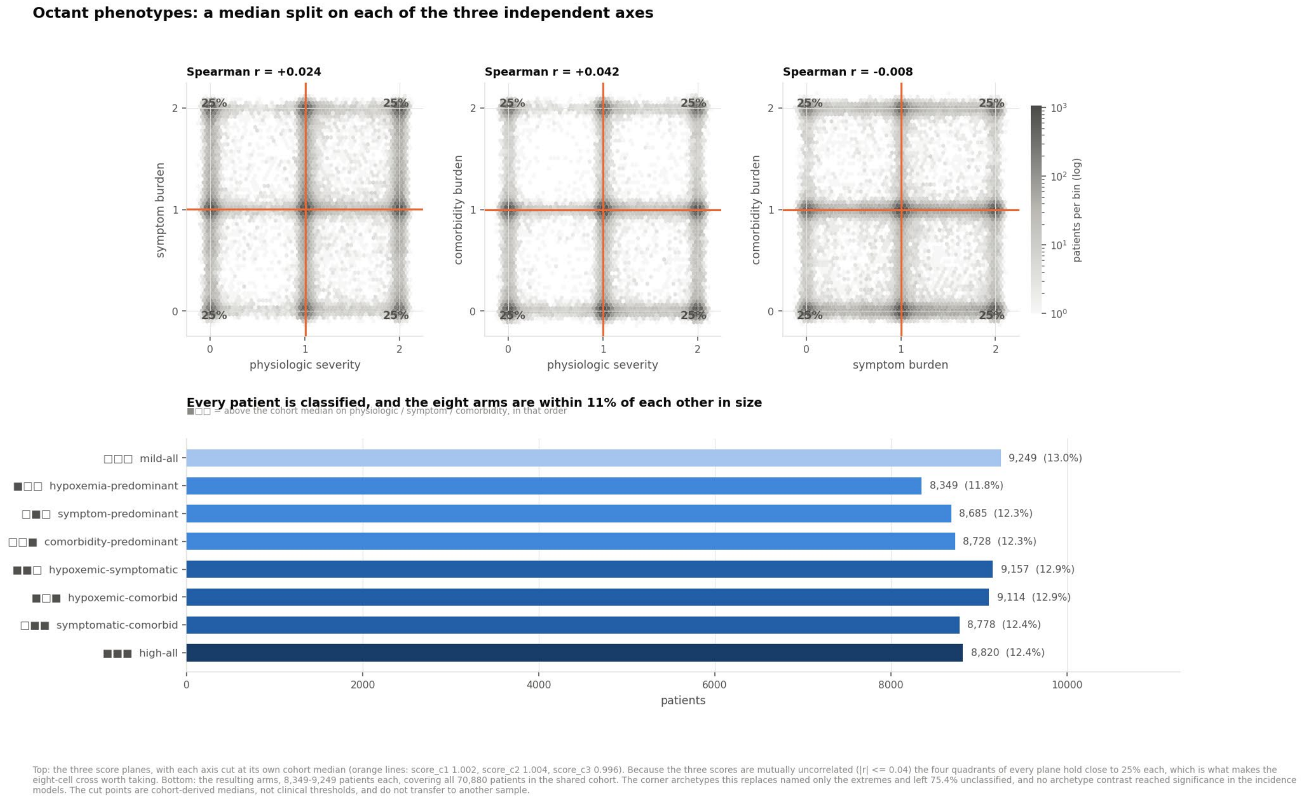


**Supplementary Figure S1**. Construction of the eight Phase 3 cross-domain phenotype octants. Posterior-weighted physiology, symptom, and comorbidity scores were split at shared-cohort medians (1.0023, 1.0041, and 0.9959). Pairwise Spearman correlations were 0.024, 0.042, and −0.008. The resulting eight strata included 8,349–9,249 participants and classified all 70,880 members of the shared phenotype cohort. The boundaries are cohort-derived descriptive cuts, not clinical thresholds.
